# Extracting Social Determinants of Health from Electronic Health Records: Development and Comparison of Rule-Based and Large Language Model Methods

**DOI:** 10.1101/2025.11.15.25339520

**Authors:** Bo Wang, Dia Kabir, Cheryl R. Clark, Karmel W. Choi, Jordan W. Smoller

## Abstract

**Background:** Social determinants of health (SDoH) are critical drivers of health outcomes but are often under-documented in structured electronic health record (EHR) data. Instead, SDoH are more commonly recorded in unstructured clinical notes, and unlocking this information could have far-reaching implications for advancing population health research and inform clinical decision making.

**Objectives:** This study aimed to develop and systematically evaluate cost-efficient methods for extracting SDoH information from unstructured clinical text using rule-based natural language processing (NLP) and large language model (LLM)-based approaches.

**Methods:** We constructed a gold-standard annotated corpus comprising clinical text segments from 171 patients in the Mass General Brigham Research Patient Data Registry, covering seven SDoH domain categories and 23 subcategories. A rule-based system (RBS) was developed and evaluated alongside seven OpenAI GPT models (GPT-4o, 4.1, 4.1-mini, o4-mini, GPT-5, GPT-5-mini, and o3) under zero-shot and few-shot settings with multiple prompting strategies. We additionally implemented late-fusion ensemble approaches that combined outputs from rule-based and LLM-based methods. Performance was assessed using precision, recall, and F1 score, alongside qualitative error analysis.

**Results:** The RBS achieved high precision for SDoH domain categories (0.96) but substantially lower recall (0.68). GPT-based models consistently outperformed RBS in overall recall and F1 scores. The best domain-level performance was observed for GPT-5 and GPT-5-mini in few-shot settings (F1=0.89), while o4-mini achieved the highest subcategory-level performance (F1=0.88). A late-fusion ensemble integrating RBS and GPT outputs further improved domain-level performance (F1=0.92), with balanced precision (0.93) and recall (0.90), but did not improve subcategory-level performance.

**Conclusion:** Recent GPT models with advanced reasoning capabilities, including the newly released “mini” models (e.g., o4-mini and GPT-5-mini), demonstrated strong performance for SDoH extraction without task-specific fine-tuning and consistently outperformed the rule-based NLP system.

Integrating rule-based and LLM-based methods via late-fusion further enhanced domain-level extraction performance. Our results demonstrate a cost-efficient framework for the accurate identification of SDoH from clinical text, facilitating downstream population health research and clinical informatics applications.

## INTRODUCTION

Social determinants of health (SDoH) are increasingly recognized as critical factors influencing health outcomes and contributing to health disparities. Unmet social needs, such as financial hardship, food insecurity, housing instability and lack of social support, are estimated to account for 30-55% of health outcomes [1]. In recent years, electronic health records (EHRs) have been a crucial resource of real-world data with applications for risk stratification, pharmacoepidemiology, treatment response prediction, and more. However, the value of EHR-based research has been limited by the under-documentation of important patient information such as SDoH within structured EHR data. Instead, this information is more commonly captured in unstructured clinical notes [2–4]. Unlocking the potential of this SDoH information could have far-reaching implications for population health research and inform clinical decision making [2].

Consequently, a growing body of work has focused on extracting SDoH from narrative clinical notes using natural language processing (NLP). Historically, these efforts have relied on either rule-based [5–8] or supervised machine learning approaches [3,9,10]. Although rule-based approaches are interpretable and customizable, they often suffer from low sensitivity due to their dependence on fixed, manually engineered rules. Conversely, supervised learning methods require significant amounts of high-quality annotated training data which can be cost- and labor-intensive to generate. This dependence on annotated data is reflected in prior studies such as the 2022 n2c2/UW shared task [20,21], in which the top performing systems used transformer-based models fine-tuned on annotated corpora. Recent advances in large language models (LLMs) and LLM ensembling [11–14] present an opportunity to develop scalable solutions for identifying SDoH without the need for substantial annotated data [15–17].

Despite progress, several important gaps remain. First, the scope of SDoH domains addressed in most studies is limited. A 2021 review by Patra et al. [2] found that smoking status is among the most commonly studied SDoH-related domains, followed by substance abuse and homelessness. In contrast, SDoH factors such as financial problems, social support, food security, and health insurance coverage, remain relatively underexplored [17,18]. Second, rule-based approaches accounted for approximately one-quarter of the reviewed studies (22 of 82) [2]. Among studies exploring the use of LLMs, most have employed either open-weight models such as LLaMA-2 and FLAN-T5 [15,17,19,20] or earlier versions of proprietary LLMs like GPT-3.5 [16,21], while more advanced LLMs with superior reasoning capabilities (including in healthcare contexts [22,23]) remain largely unexplored for SDoH extraction. Notably, a recent study by Keloth et al. [17] emphasized the importance of evaluating the latest LLMs for this task. Lastly, while fine-tuning open-weight models has gained traction [15,17,19,21], the development and systematic evaluation of different prompting strategies with state-of-the-art LLMs, including fusion with rule-based systems, remains understudied despite its relevance in resource-constrained settings.

Our study addresses the aforementioned gaps by developing and evaluating methods to identify seven SDoH domains from clinical text derived from multiple note types. In addition to commonly studied domains, we emphasize less-explored determinants such as social resources and health insurance status, as well as physical activity, a key behavioral determinant of health. A recent study by Lituiev et al. [24] used a two-tier taxonomy with first- and second-level SDoH classes. Similarly, we developed a fine-grained classification system for each domain. For example, within health insurance status, we included subcategories such as *adequate insurance coverage*, *lack of insurance* and *government-assisted insurance* to capture different levels of access to care and coverage. Inspired by the n2c2/UW shared task [25], we further annotated each subcategory with four contextual attributes (*temporality*, *experiencer*, *hypothetical status* and *uncertainty*). These attributes served as exclusion criteria for defining positive SDoH cases and supported qualitative analysis of the models’ handling of contextual distinctions. This annotation framework was designed to capture richer social context information for downstream applications such as health risk stratification.

We present two complementary NLP approaches for SDoH extraction requiring minimal training and computational resources: a rule-based system (RBS) and LLM-based methods leveraging state-of-the-art OpenAI Generative Pretrained Transformer (GPT) models at the time of experimentation. For the LLM-based approach, rule-based pre-screening was used to identify candidate text segments from clinical notes, which were then classified by GPT models. We examined seven GPT models in both zero-shot and few-shot settings without task-specific fine-tuning, evaluating performance at the segment-level. Our prompting strategies incorporated examples that varied in annotation difficulty. This work extends prior studies by systematically evaluating advanced reasoning models and comparing their performance against RBS. Finally, we investigated various ensemble strategies using late fusion to combine both approaches and further improve extraction performance. Overall, this study contributes to addressing key gaps in SDoH research and provides a cost-efficient framework for future clinical NLP applications.

## METHODS

### Ethical considerations

All procedures contributing to this work comply with the ethical standards of the relevant national and institutional committees on human experimentation and with the Helsinki Declaration of 1975, as revised in 2008. The study protocol (2018P002642) was approved by the MGB Institutional Review Board, with an informed consent waiver for the use of retrospective medical record data without patient interaction. Although the clinical notes used in this study were not deidentified, all data access, modeling, and analysis were restricted to authorized researchers and conducted within secure environments behind the MGB firewall. No identifiable patient information is included in the manuscript or supplementary materials.

### Data source

Data were obtained from the Mass General Brigham (MGB) Research Patient Data Registry (RPDR) [26], a centralized data registry of clinical information from EHRs across the MGB health system. The RPDR database includes approximately seven million patients from eight hospitals, including two major teaching hospitals, Massachusetts General Hospital and Brigham and Women’s Hospital, encompassing a wide range of patient characteristics including structured data (demographics, diagnoses, medications, procedures, and lab tests) and unstructured narrative notes. We acquired all narrative clinical notes from 95,157 patients enrolled in the MGB Biobank [27], with notes spanning March 1976 to November 2021. For this study, we used discharge summaries and progress notes, the latter including inpatient, outpatient, and emergency visit notes.

### SDoH categories

We established our SDoH classification schema (Figure 1) through iterative consultations with subject matter experts in psychology, psychiatry, and health disparities research. The final schema encompasses seven domains, i.e., six social and one behavioral (physical activity) determinants of health commonly screened in clinical practice [28–30]. To capture granular social contexts relevant to health outcomes, we further defined multiple subcategories within each domain category. Given the inherent overlap between certain subcategories (e.g., “patient recently got laid off” could be characterized as both *job loss* and *unemployment*), we developed comprehensive annotation guidelines (Appendix S2) to ensure consistency and reproducibility in data generation. The schema was also refined to draw more nuanced distinctions. For example, within *Social Resources*, we distinguished subcategories of *Living with or accompanied by someone* and *living alone* from *good social resources* and *poor social resources* to separate living situation from quality of social support. The initial taxonomy of 27 subcategories was refined to 23 based on data review and feasibility assessment. Instances falling within a domain category but not matching any specific subcategory were assigned to “NA” (not applicable).

**Figure 1.**
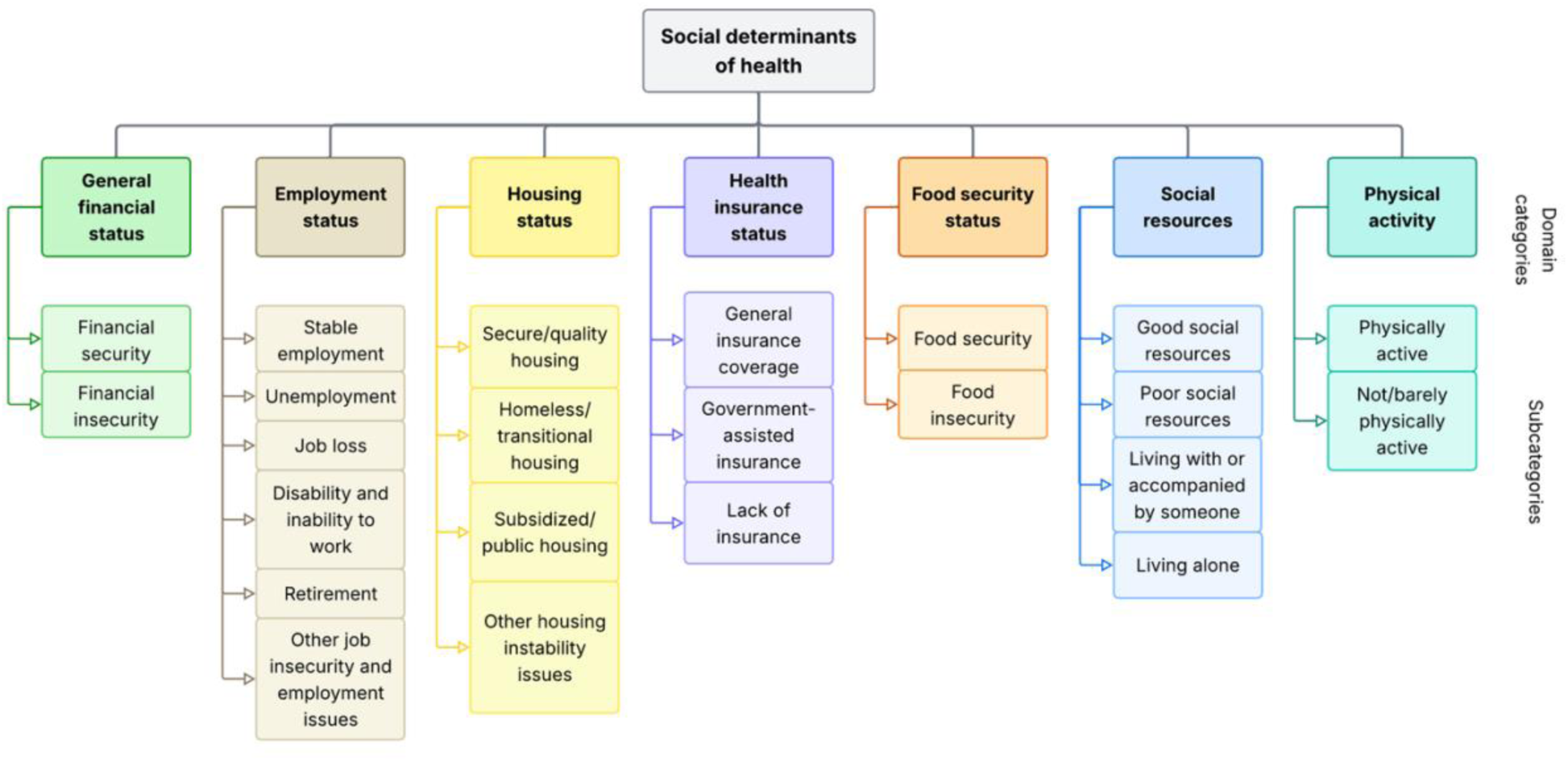
SDoH classification schema, comprising seven domains: General financial status (green), Employment status (beige), Housing status (yellow), Health insurance status (purple), Food security status (orange), Social resources (blue), and Physical activity (teal). Domain categories are shown in bold, with their corresponding subcategories listed below each domain (23 subcategories in total).

### Gold-standard corpus development

An initial set of 2,000 patients was sampled from the MGB Biobank cohort using stratified sampling based on several key sociodemographic variables: sex, self-reported race, age group (≥65 vs. <65), and health insurance type (public vs. private payer). This cohort was held out from system development, including lexicon refinement of RBS, to ensure separation from evaluation. To create a gold-standard dataset, we conducted pre-screening using predefined patterns from our rule-based system (see next section), including keywords, rules, and regular expression-based matching, to identify sentences in patients’ notes likely containing SDoH information.

Following prior work [9,31], for each potential SDoH mention, we extracted text segments with 150 characters of context on either side, providing sufficient information for annotation. We randomly sampled 79 unique text segments for annotator training, and a separate set of 226 text segments (10 per subcategory, except “*Other job insecurity and employment issues*” which had 6 instances) for primary model validation. These 226 segments were drawn from notes with a median documentation year of 2016 (IQR: 2012-2019).

We developed our annotation workflow using Label Studio (https://labelstud.io/), an open-source data labeling platform (Figure S1). Two annotators (BW: a postdoctoral researcher trained in biomedical informatics; DK: a medical student with experience in neuropsychiatric research) underwent three training sessions and calibration meetings before final annotation. Disagreements between annotators were discussed and resolved in consensus meetings with a clinical psychologist (KWC) serving as the tiebreaker. Based on the finalized SDoH classification schema (Figure 1) and annotation guidelines, we defined two multi-label annotation tasks for each text segment:

- Task 1: Identify mentions of the 7 SDoH domain categories.
- Task 2: Identify mentions of the 23 SDoH subcategories.

Each training session began with detailed discussion of the annotation guidelines and concluded with inter-annotator agreement (IAA) assessment. After completing three training sessions, annotators achieved acceptable IAA as measured by Krippendorff’s alpha [32] (α = 0.88 for domain categories, α = 0.74 for subcategories). They then independently annotated the final 226 text segments, achieving α = 0.95 for domain categories, 0.84 for subcategories alone, and 0.78 for subcategories with contextual attributes (temporality, experiencer, hypothetical status, and uncertainty), all well above standard thresholds for reliability [33].

### Rule-based system

As illustrated in Figure 2, we developed a rule-based system (RBS) to identify SDoH mentions in clinical notes. The system was implemented using medspaCy [34], an open-source clinical NLP library that supports flexible integration of rule-based and machine learning-based algorithms. It employs a pipeline of NLP functions spanning sentence and section detection to concept extraction and contextual analysis, to locate, match and disambiguate SDoH mentions, and an iterative process of manual testing, review and refinement based on feedback to optimize model performance. Descriptions for each pipeline component are provided in Appendix S1.3.

**Figure 2.**
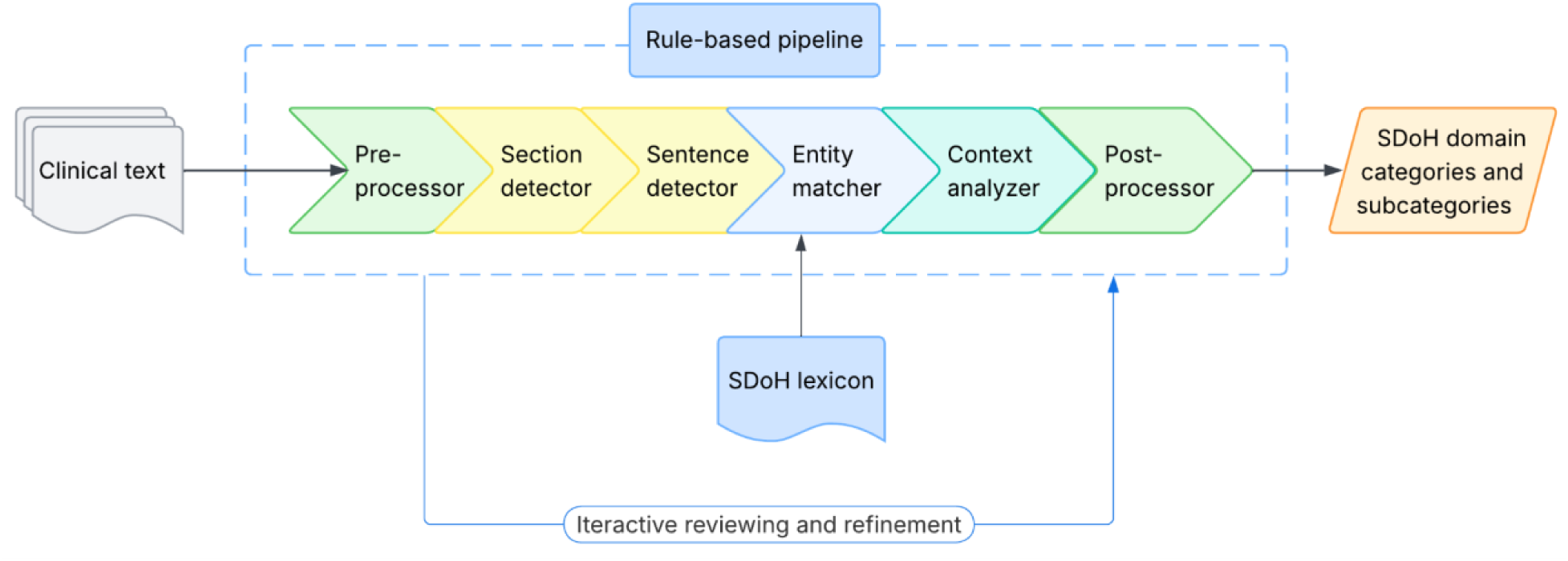
RBS workflow, which takes clinical notes as input, processes them through its text processing pipeline, including sentence segmentation, lexicon-based entity matching, and context disambiguation, and outputs sentences identified with SDoH domain categories and subcategories. The pipeline was developed iteratively involving manual reviewing and refinement of the rules.

### Lexicon creation and expansion

RBS requires a lexicon (a dictionary of relevant query terms) and corresponding rules for identifying matches in clinical text. We developed our SDoH lexicon through a multi-step process: seed term curation, lexicon expansion, and iterative filtering and refinement (Figure 3). The initial lexicon comprised terms related to each domain derived from prior studies and reviews [6,8,29,30,35–39] and SDoH screening tools and surveys (see Appendix S1.1). These seed terms were reviewed by a domain expert (CRC) and systematically expanded using: (1) synonym identification and hierarchy mapping (hypernymy-hyponymy) through UMLS Metathesaurus, and (2) semantic similarity search using text embedding models [40–45]. For instance, we encoded the MEDLINE N-Gram Set (1-5 grams) [46,47] using an MPNet-based Sentence-BERT model (*paraphrase-mpnet-base-v2*) [44] (which we make publicly available at: https://github.com/bwang482/SDoH_Extraction) and performed semantic searches using seed terms as queries against the embedded corpus to identify additional relevant SDoH terms. The iterative lexicon filtering step is detailed in Appendix S1.2.

**Figure 3.**
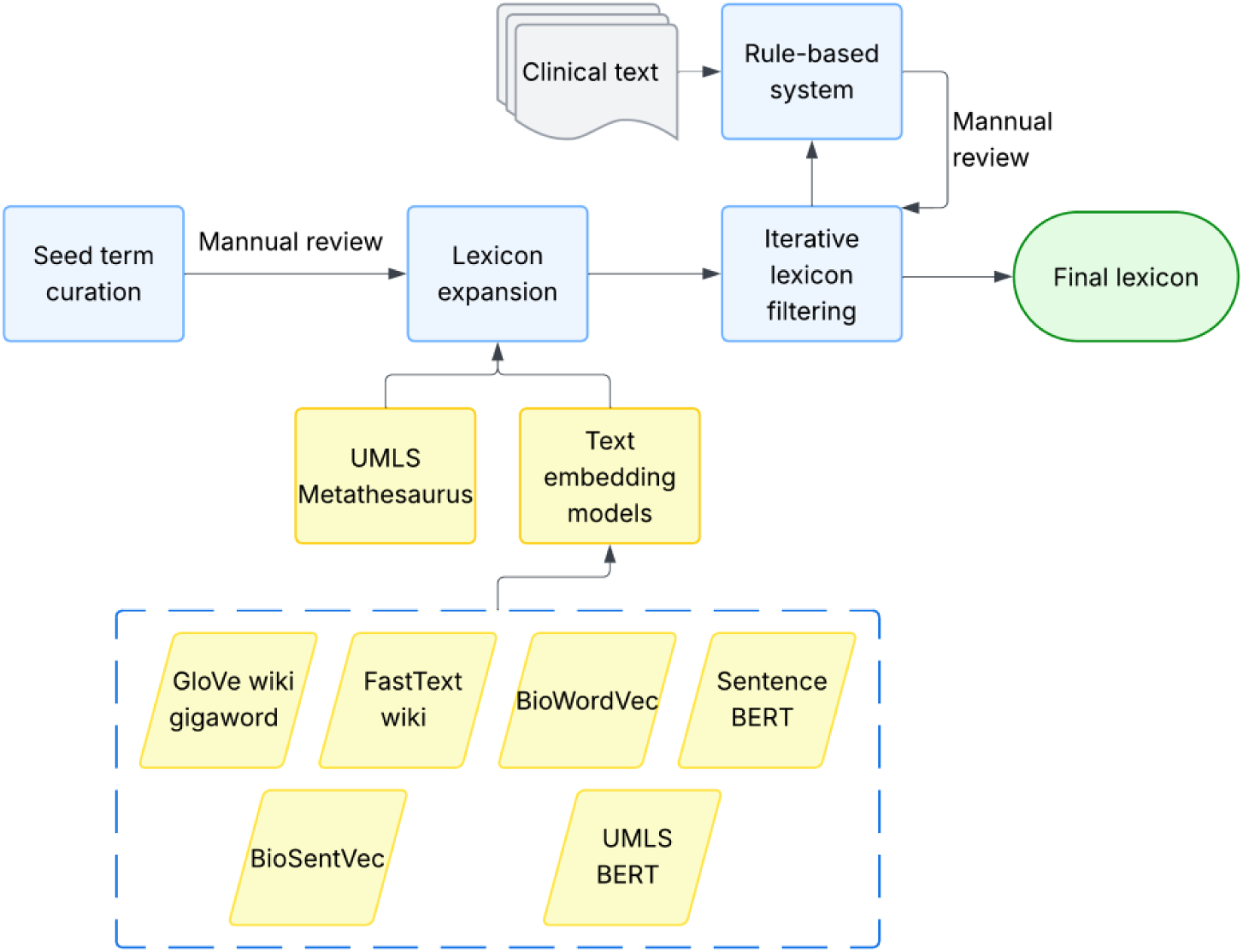
Lexicon curation process, involving seed term curation, lexicon expansion using UMLS and embedding-based semantic search, and an iterative process of lexicon filtering.

### LLM-based models

We employed a series of OpenAI GPT models via the Azure OpenAI Service (Microsoft Foundry), deployed within MGB’s HIPAA-compliant Microsoft Azure infrastructure. Using the Chat Completions API, we prompted the models to identify SDoH categories and subcategories in clinical text segments, akin to a hierarchical, multi-label classification task.

While open-weight LLMs continue to improve, frontier proprietary models remain state-of-the-art on many benchmarks, particularly those involving reasoning tasks [23,48,49]. For this study, we selected seven GPT models for SDoH extraction based on their performance in benchmarks including HealthBench [22], as well as their inference cost: GPT-4o, 4.1, 4.1-mini, 5, 5-mini, o3, and o4-mini (model versions are provided in Appendix S4). For reasoning models (i.e., GPT-5, 5-mini, o3, and o4-mini), the reasoning effort parameter was set to high. We compared their precision and recall with those of the RBS, evaluated performance (in precision, recall and F1 scores) and conducted error analysis to understand model limitations and performance variability.

### Zero-shot prompting

State-of-the-art LLMs are instruction-tuned on diverse tasks [50,51], enabling them to follow novel instructions and perform unseen tasks without requiring additional training, a capability known as zero-shot generalization [52,53]. Prior studies have highlighted the importance of systematic prompt engineering [17,20,54], for example by incorporating annotation guideline instructions. As shown in Figure S2 and Appendix S3, our zero-shot approach employed a three-component system prompt: (1) role-playing instructions to contextualize the task, (2) definitions of SDoH domain categories and subcategories, and (3) step-by-step instructions for extracting SDoH information and structuring the output. The prompt instructed LLMs to follow a hierarchical approach: identify SDoH domain categories first, then their corresponding subcategories, returning “None” if no relevant information was present. We emphasized that only current, patient-specific, and confirmed SDoH information should be extracted, excluding historical, family-member, or hypothetical mentions (e.g., doctor’s recommendations).

In addition, we evaluated three prompting styles, namely strict, balanced, and liberal, which differ in their threshold for identifying SDoH domains and subcategories (inclusion criteria). The *strict* style requires explicit evidence, *balanced* accepts evidence that is strongly implied, and *liberal* takes the most inclusive approach, accepting not only explicit and strongly implied evidence but also information deemed reasonably likely from the context. We hypothesized that these styles would demonstrate a precision-recall trade-off, with *strict* achieving higher precision and *liberal* yielding higher recall.

### Few-shot prompting

Following [16], we tested six few-shot prompting strategies using examples from our annotation training: (1) using five examples where annotators readily agreed on the correct identification of SDoH information (i.e., easy examples); (2) same as (1), but with explanations describing the reasoning behind each annotation; (3) using five challenging examples that required annotator adjudication (i.e., hard examples); (4) same as (3), but with explanations added; (5) using synthetically generated examples by GPT; (6) same as (5), but with synthetic explanations. The goal of these experiments was to assess whether advanced GPT models benefit from examples with or without explanations, and whether exposure to difficult, ambiguous, or synthetic cases, improves their accuracy and robustness in SDoH extraction. The few-shot prompt structure is illustrated in Figure S3.

### RBS-GPT ensemble

Drawing on prior work in LLM ensemble learning within the broader NLP domain [11–14], we implemented a hybrid ensemble approach that aggregates rule-based and GPT model outputs using late fusion to improve the overall performance of SDoH extraction. Using the annotator training samples as development data, we found RBS achieved good precision (0.92) for domain categories but showed poor performance (precision = 0.64) at the subcategory level. Based on these findings, we designed two different ensemble strategies (Figure 4): (a) for domain categories, we deployed the best performing GPT model with three prompting styles (*strict*, *balanced*, and *liberal*) to form a small “GPT-committee”. The committee’s outputs were first aggregated and then combined with those from RBS; (b) for subcategories, we used only the aggregated GPT-committee output to determine the final list of SDoH subcategories. We evaluated several ensemble configurations, testing three fusion functions for the GPT-committee: (1) majority voting, (2) union, and (3) intersection, and compared them with GPT using strict prompting alone. We also examined two fusion functions (union and intersection) for combining RBS and GPT-committee outputs (RBS-GPT fusion).

**Figure 4.**
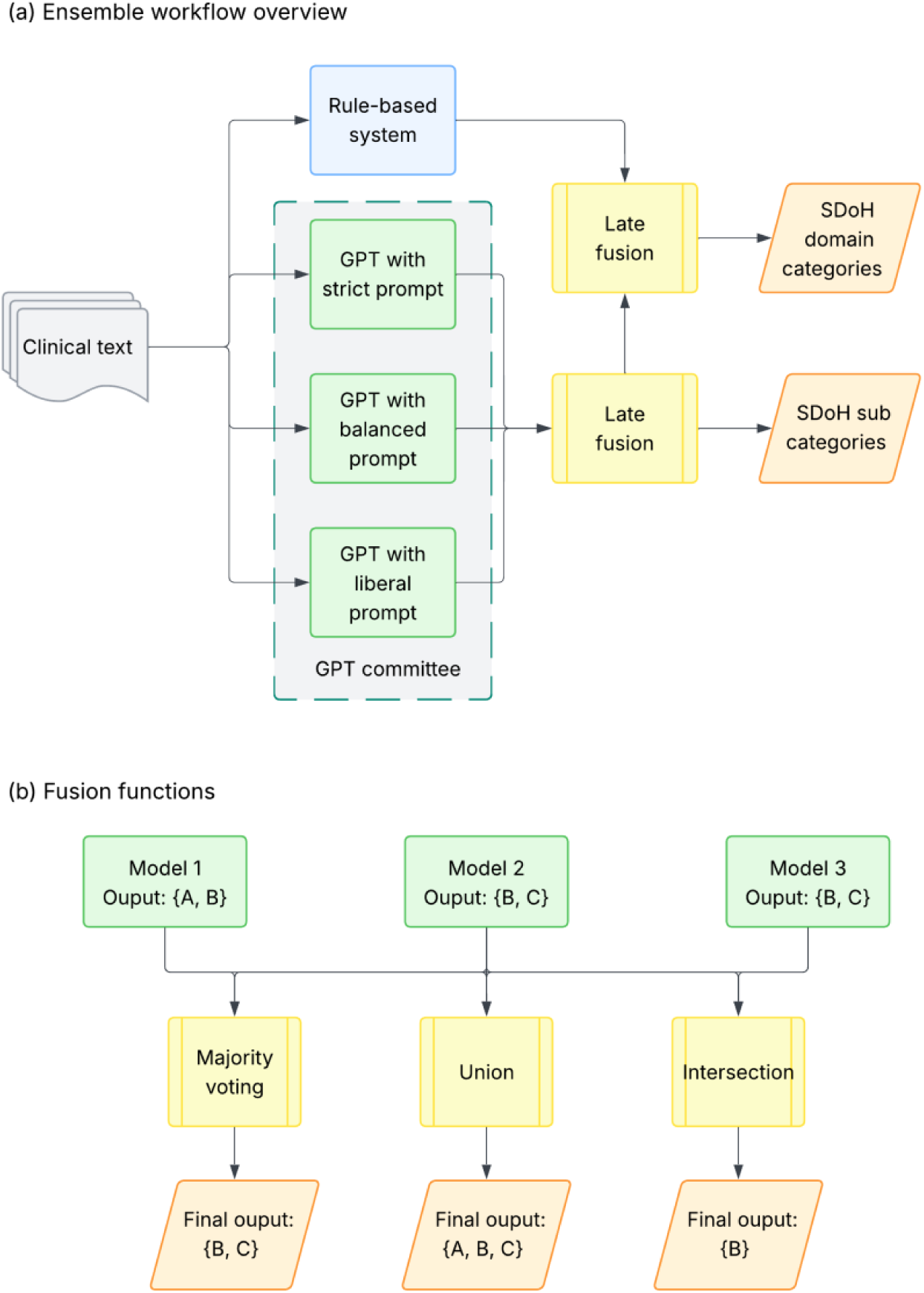
(a) Overview of the RBS–GPT ensemble workflow, showing separate fusion paths for SDoH domain categories and subcategories. For domain categories, outputs from both RBS and GPT with different prompting styles (the GPT-committee) are aggregated via late fusion. For subcategories, only the GPT-committee outputs are aggregated. (b) Illustrative example of the fusion functions, namely *majority voting*, *union* and *intersection*.

## RESULTS

### Gold-standard data annotation

The final annotated validation dataset consisted of 226 text segments from 171 patients, with a total of 410 SDoH domain-category annotations, including 7 labeled “None of the Above” (NOA), and 475 subcategory annotations, including 153 labeled “NA”. Table 1 summarizes patient demographic characteristics, showing balanced distributions of gender and self-reported race, with a predominance of older patients and those with public health insurance.

**Table 1.**
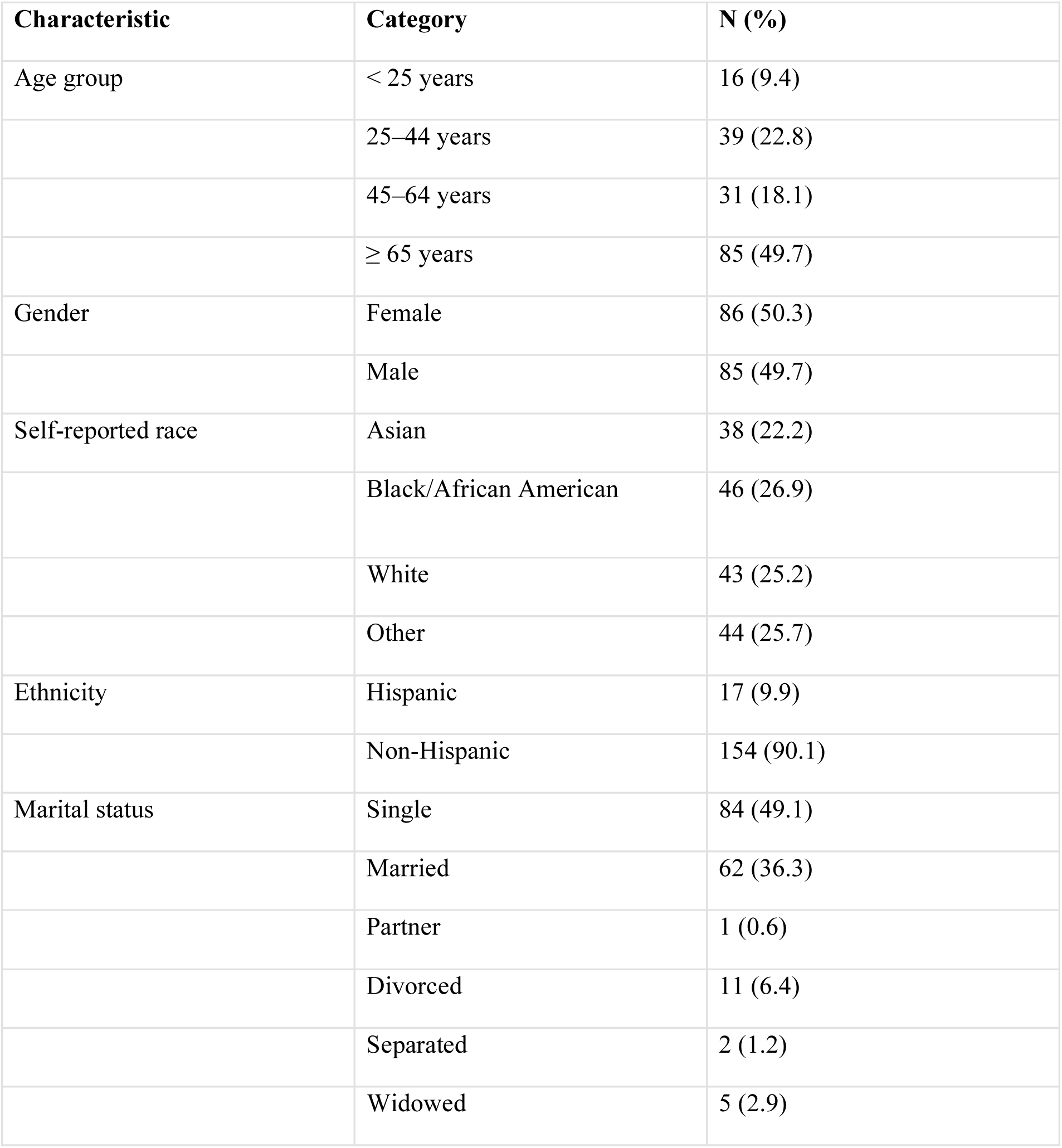
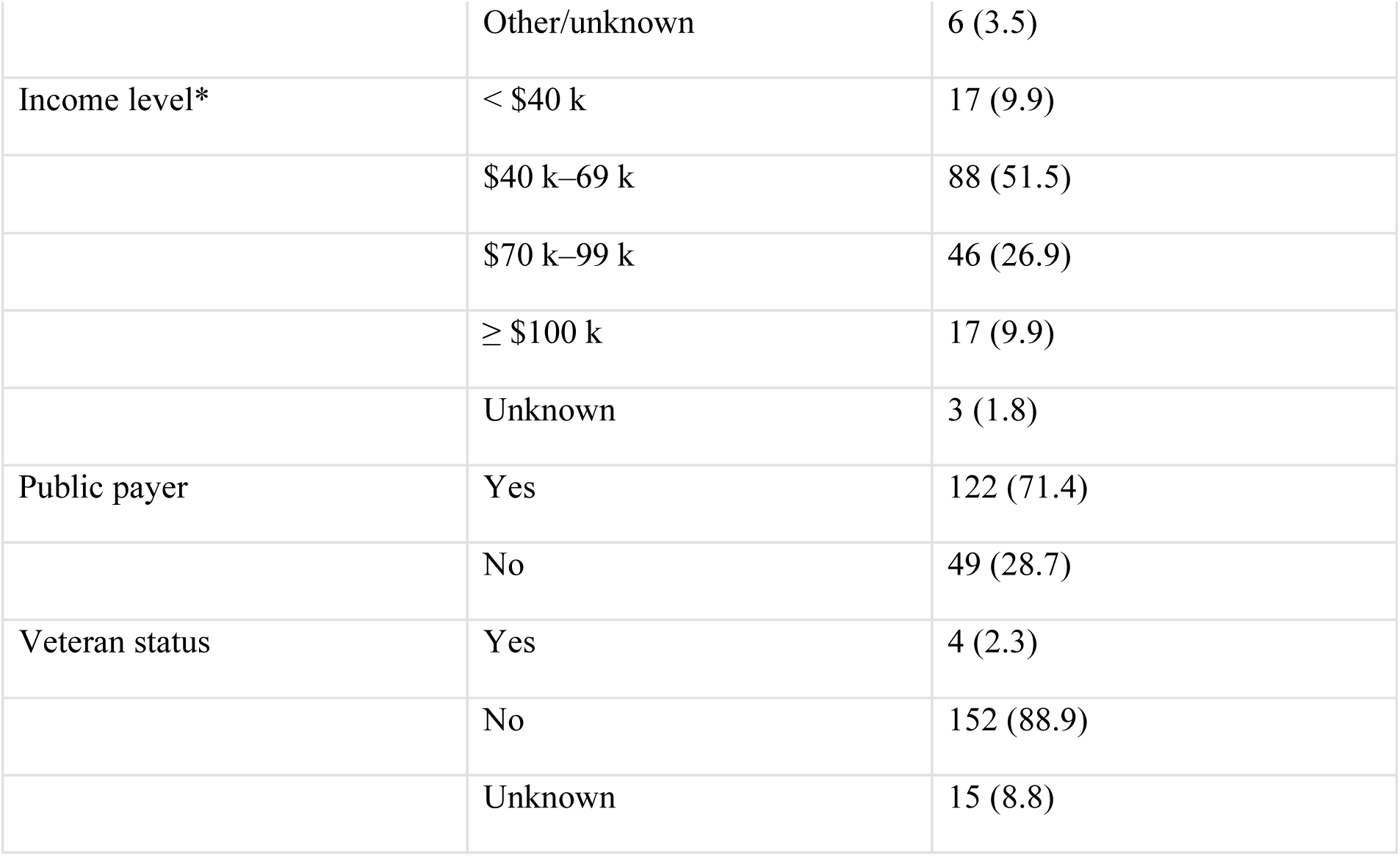
Patient demographics from the annotated validation dataset. *: Median household income by ZIP code.

| Characteristic | Category | N (%) |
| --- | --- | --- |
| Age group | < 25 years | 16 (9.4) |
|  | 25–44 years | 39 (22.8) |
|  | 45–64 years | 31 (18.1) |
|  | ≥ 65 years | 85 (49.7) |
| Gender | Female | 86 (50.3) |
|  | Male | 85 (49.7) |
| Self-reported race | Asian | 38 (22.2) |
|  | Black/African American | 46 (26.9) |
|  | White | 43 (25.2) |
|  | Other | 44 (25.7) |
| Ethnicity | Hispanic | 17 (9.9) |
|  | Non-Hispanic | 154 (90.1) |
| Marital status | Single | 84 (49.1) |
|  | Married | 62 (36.3) |
|  | Partner | 1 (0.6) |
|  | Divorced | 11 (6.4) |
|  | Separated | 2 (1.2) |
|  | Widowed | 5 (2.9) |
|  | Other/unknown | 6 (3.5) |
| Income level* | < \$40 k | 17 (9.9) |
| | \$40 k–69 k | 88 (51.5) |
| | \$70 k–99 k | 46 (26.9) |
| | ≥ \$100 k | 17 (9.9) |
|  | Unknown | 3 (1.8) |
| Public payer | Yes | 122 (71.4) |
|  | No | 49 (28.7) |
| Veteran status | Yes | 4 (2.3) |
|  | No | 152 (88.9) |
|  | Unknown | 15 (8.8) |

As noted earlier, the two annotators achieved strong inter-annotator agreement (Krippendorff’s α: 0.95 for domain categories, and 0.78 for subcategories with attributes). The majority (64%) of annotation disagreements for domain categories were attributed to “Social resources” (which required clearer scope definition) and “General financial status” (where one annotator overlooked questionnaire-derived mentions). Similarly, most subcategory-level disagreements stemmed from poorly formatted in-text questionnaires and inconsistencies in identifying contextual attributes of SDoH mentions. For instance, references to public housing applications prompted discussion about whether they should be labeled as “Hypothetical” under “Subsidized/public housing”; additionally, annotators needed further guidance to correctly parse question-answer pairs within poorly formatted questionnaire text. Only text segments containing current, patient-specific, and non-hypothetical SDoH information were considered as SDoH-positive cases.

### Model performance

Figure 5 summarizes the macro-averaged performance of the rule-based system (RBS) and five GPT-based models (additional results in Table S1); 95% confidence intervals for macro- and micro-averaged F1 scores are shown in Figure S4 and S5. While RBS achieved high precision for domain categories, it demonstrated substantially lower recall compared to GPT-based models across both classification levels, highlighting the inherent limitation of fixed rules. In contrast, GPT-based models consistently outperformed RBS in recall and F1 scores. At the domain-category level, GPT-5-mini (5-shot) and GPT-5 (5-shot) were tied for the highest point-estimate F1 score (0.89), and precision/recall/F1 values of 0.91/0.87/0.89 (95% CI for F1, 0.86-0.91) and 0.96/0.84/0.89 (95% CI for F1, 0.86-0.92), respectively. At the subcategory level, the o4-mini models demonstrated the strongest performance, achieving an F1 of 0.87 (95% CI, 0.83-0.90) in the zero-shot setting (precision/recall/F1: 0.85/0.91/0.87) and 0.88 (95% CI, 0.84-0.91) in the 5-shot setting (0.90/0.87/0.88). As shown in Figure S5, the 95% confidence intervals overlapped considerably between o4-mini and GPT-5, suggesting that small differences in point estimates should be interpreted cautiously. Overall, few-shot prompting yielded modest gains in precision for subcategories, often accompanied by slight reductions in recall. Table 2 reports micro-averaged metrics and absolute error counts (FP, FN) for subcategories (additional results in Table S2). Consistent with the macro-averaged results, o4-mini remained the best overall subcategory model, with o4-mini (5-shot) achieving the highest micro-averaged F1 (0.88; 95% CI, 0.85-0.91; TP/FP/FN = 283/37/39). GPT-5-mini (zero-shot) achieved the highest recall (0.93) and the largest number of true positives (TP = 298), but at the cost of more false positives (FP = 100).

**Figure 5.**
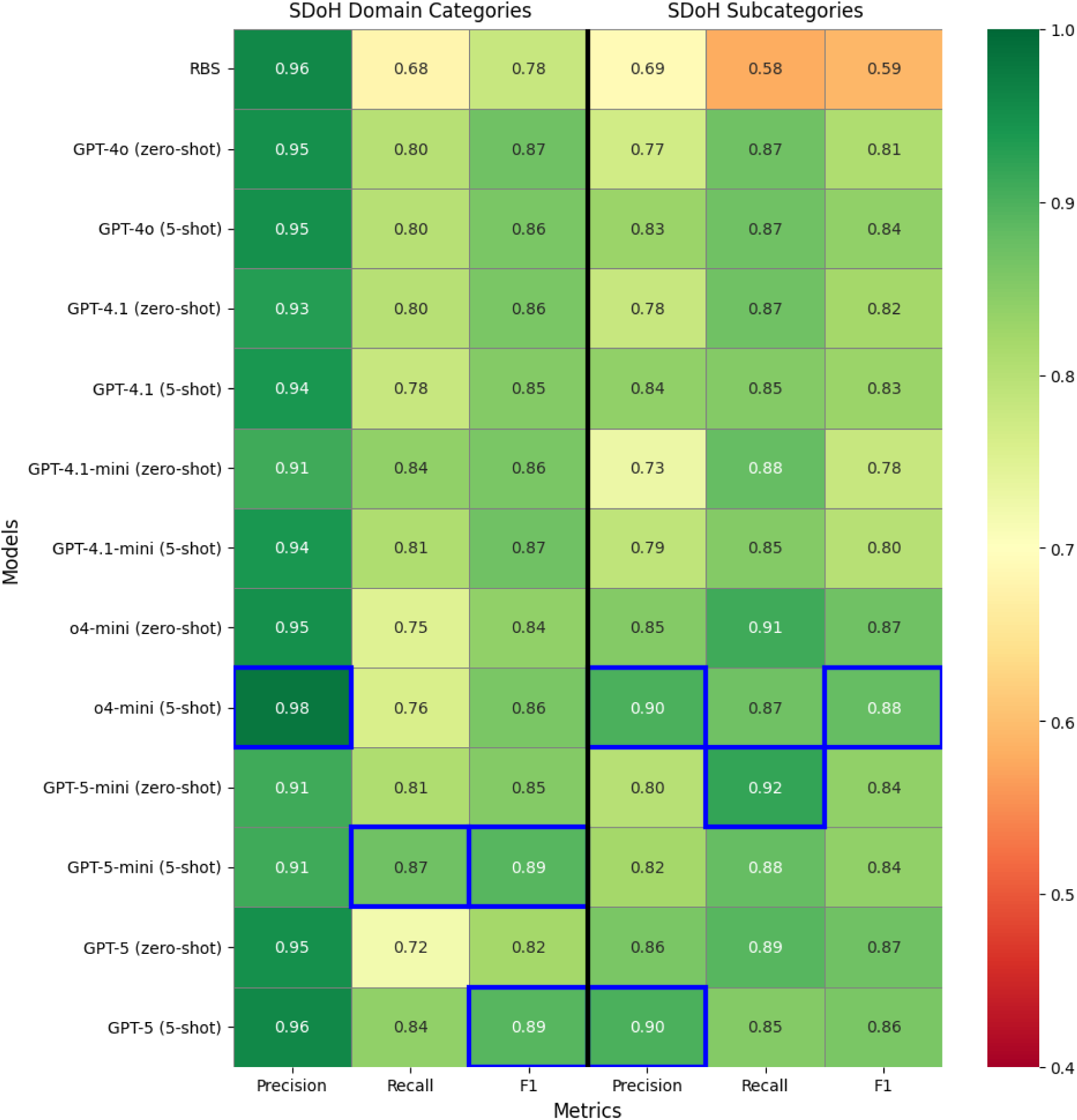
Overall performance comparison between RBS and GPT-based approaches. Macro-averaged precision, recall, and F1 are reported separately for domain categories and subcategories. Corresponding 95% confidence intervals for macro-F1 are presented in Figure S4 and S5. All reasoning models reported here, namely o4-mini, GPT-5-mini, and GPT-5, were run with the high-reasoning setting. The highest score for each metric (column) is highlighted with a blue box border.

**Table 2.** Micro-averaged performance metrics and absolute error counts for subcategories. Absolute counts denote true positives (TP), false positives (FP), and false negatives (FN) in relative to gold-standard annotations. Note: TP + FN sums to 322 rather than the 475 total annotations because 153 annotations labeled “NA” do not contribute to TP or FN for any positive subcategory label.

| Models | Precision | Recall | F1 | TP | FP | FN |
| --- | --- | --- | --- | --- | --- | --- |
| Rule-based (RBS) | 0.67 | 0.49 | 0.57 | 159 | 79 | 163 |
| GPT-4o (zero-shot) | 0.77 | 0.85 | 0.81 | 274 | 83 | 48 |
| GPT-4o (5-shot) | 0.82 | 0.84 | 0.83 | 272 | 58 | 50 |
| GPT-4.1 (zero-shot) | 0.79 | 0.85 | 0.82 | 275 | 75 | 47 |
| GPT-4.1 (5-shot) | 0.84 | 0.83 | 0.83 | 266 | 50 | 56 |
| o4-mini (zero-shot) | 0.84 | 0.90 | 0.87 | 289 | 55 | 33 |
| o4-mini (5-shot) | 0.88 | 0.88 | 0.88 | 283 | 37 | 39 |
| GPT-5-mini (zero-shot) | 0.75 | 0.93 | 0.83 | 298 | 100 | 24 |
| GPT-5-mini (5-shot) | 0.79 | 0.90 | 0.84 | 289 | 78 | 33 |
| GPT-5 (zero-shot) | 0.84 | 0.88 | 0.86 | 282 | 54 | 40 |
| GPT-5 (5-shot) | 0.88 | 0.84 | 0.86 | 272 | 37 | 50 |

Appendix S5 benchmarks the inference time (client wall-clock) and cost across o4-mini, GPT-5-mini and GPT-5 under a high-reasoning setting, using a single-worker sequential deployment. While both the mini models performed competitively with GPT-5, they demonstrated substantially lower inference latency and estimated per-segment cost. For example, o4-mini required approximately 7.0 seconds per segment in zero-shot and 8.9 seconds in 5-shot prompting, compared with 33.0 and 34.6 seconds, respectively, for GPT-5. API-reported reasoning tokens accounted for most completion tokens across models (94-98%), reflecting the computational overhead of chain-of-thought processing. Based on cost-efficiency and competitive performance at both levels, as demonstrated in the per-domain and per-subcategory results (Table S3), we selected o4-mini (5-shot) as our primary model for further experiments involving ensemble modeling.

### Prompting comparison

Among the strict, balanced, and liberal prompting styles, we observed the anticipated precision-recall trade-off (Table S4): strict prompting achieved the highest precision and F1 scores for subcategories in both zero-shot and 5-shot settings, while liberal prompting had the best recall, and balanced prompting yielded intermediate results.

Performance varied modestly across example types (easy, hard, or synthetic), with F1 scores ranging from 0.81-0.86 for domain categories and 0.87-0.88 for subcategories (Table S5). Hard examples requiring annotator adjudication yielded better performance for subcategories, particularly in precision (0.90 vs. 0.86 for easy examples and 0.88 for synthetic examples). Adding explanations to the prompt improved precision for subcategories in the hard-example model (from 0.88 to 0.90) but had little impact on other models or metrics.

### Ensemble performance

As described in Methods, we used different ensemble strategies by classification level: two-step fusion for domain categories and GPT-committee ensembling for subcategories. As shown in Table 3, *majority voting* on GPT-committee outputs followed by *union* (∪) with RBS outputs achieved the highest macro-averaged F1 (0.92; 95% CI, 0.89-0.94) and the most balanced performance for domain categories, with precision above 0.93 and recall above 0.90. However, the overlapping confidence intervals suggest that performance differences among top fusion strategies should be interpreted cautiously. Of the 402 domain-category predictions produced (of which 21 were NOA), 32 (8.0%) were contributed by RBS alone, demonstrating its added value; 140 (34.8%) came from the GPT-committee, indicating their ability to capture patterns missed by RBS; and the remaining 230 (57.2%) were identified by both systems. For subcategories, the *Strict-only* configuration achieved the best performance (precision = 0.90, recall = 0.87, F1 = 0.88; 95% CI, 0.84-0.91), consistent with our objective of maximizing F1 with balanced precision and recall. The *Intersection* configuration followed closely, achieving an F1 score of 0.87, with higher precision (0.92) but lower recall (0.84). Micro-averaged results and absolute error counts are reported in Table S15 and showed a similar overall pattern.

**Table 3.** Performance of ensemble models across fusion strategies. Macro-averaged precision, recall, and F1 scores are reported for domain categories and subcategories. *Majority voting*, *union*, and *intersection* denote label assignment by most, any, or all GPT committee members, respectively. *Strict-only* uses the strict-prompt GPT output without aggregation. For domain categories, ∪ denotes assignment by either RBS or the GPT committee, and ∩ denotes assignment by both. 95% confidence intervals were computed using 2,000 patient-level cluster bootstrap resamples. The highest scores for each metric (column) are shown in bold and underlined.

| GPT-committee → RBS-GPT Fusion | SDoH domain categories | | |
| --- | --- | --- | --- |
|  | Precision | Recall | F1 |
| Majority voting $\rightarrow \cup$ | 0.93 [0.90, 0.96] | 0.90 [0.87, 0.93] | **<u>0.92</u>** [0.89, 0.94] |
| Majority voting $\rightarrow \cap$ | **<u>0.99</u>** [0.98, 1.00] | 0.60 [0.55, 0.66] | 0.74 [0.70, 0.78] |
| Union $\rightarrow \cup$ | 0.89 [0.85, 0.92] | **<u>0.93</u>** [0.90, 0.96] | 0.91 [0.88, 0.93] |
| Union $\rightarrow \cap$ | **<u>0.99</u>** [0.97, 1.00] | 0.63 [0.58, 0.68] | 0.76 [0.72, 0.80] |
| Intersection $\rightarrow \cup$ | 0.95 [0.92, 0.98] | 0.85 [0.81, 0.89] | 0.90 [0.87, 0.92] |
| Intersection $\rightarrow \cap$ | **<u>0.99</u>** [0.98, 1.00] | 0.55 [0.49, 0.60] | 0.70 [0.65, 0.74] |
| Strict-only $\rightarrow \cup$ | 0.95 [0.92, 0.98] | 0.88 [0.84, 0.91] | 0.91 [0.88, 0.93] |
| Strict-only $\rightarrow \cap$ | **<u>0.99</u>** [0.98, 1.00] | 0.56 [0.51, 0.62] | 0.71 [0.67, 0.76] |
| GPT fusion | SDoH subcategories |  |  |
|  | Precision | Recall | F1 |
| Majority voting | 0.85 [0.82, 0.89] | 0.91 [0.87, 0.94] | 0.87 [0.84, 0.90] |
| Union | 0.73 [0.69, 0.78] | **<u>0.95</u>** [0.92, 0.97] | 0.82 [0.78, 0.84] |
| Intersection | **<u>0.92</u>** [0.89, 0.95] | 0.84 [0.79, 0.88] | 0.87 [0.83, 0.90] |
| Strict-only | 0.90 [0.87, 0.94] | 0.87 [0.83, 0.91] | **<u>0.88</u>** [0.84, 0.91] |

### Comparison to ICD codes

Both RBS and GPT models identified substantially more SDoH information than documentation based on ICD codes (V and Z codes). Of the 226 annotated samples, our RBS identified SDoH information in 225, and the 5-shot GPT models identified SDoH in 194 (o4-mini), 219 (GPT-5-mini), and 206 (GPT-5) cases. In comparison, relevant ICD codes were present in far fewer visits across all three time windows examined: using broad Z and V codes (see Table S12), matches were found in 84 visits within ±7 days of the corresponding clinical documentation, 128 visits within a 6-month lookback window, and 133 visits within a 12-month lookback window. Using SDoH category-specific Z and V codes (see Table S13), matches were found in only 11, 12, and 14 visits for the same time windows, respectively. These findings highlight the value of extracting SDoH from clinical text, as structured ICD coding substantially undercaptures SDoH information even when extended lookback periods are used.

### Error analysis

To better understand model limitations beyond quantitative performance metrics, we conducted a qualitative review of false positive and false negative predictions from the o4-mini (5-shot) model and the RBS at both the domain and subcategory levels. Due to its lack of semantic understanding, the RBS failed to capture many SDoH mentions that fell outside its predefined keyword rules. For example, it was unable to recognize that “patient owns multiple homes” indicates secure housing. Its finite vocabulary coverage also meant that domain-specific terms such as “Section 8” (a government-funded rent subsidy program) went unrecognized. The majority of RBS’s false positives stemmed from its failure to correctly recognize negations, temporality, hypotheticals, and experiencer attribution, which were annotated as contextual attributes in our validation data, as well as its inability to perform semantic disambiguation. Table S14 summarizes the primary error types for both approaches.

In contrast to RBS, o4-mini demonstrated its ability to interpret semantic meaning and generalize beyond keyword matching. However, it exhibited a different set of errors related to reasoning calibration, evidence evaluation, and instruction adherence. These errors can be classified into four primary categories: 1) Missing implicit evidence. Under the strict prompting style adopted for subsequent analyses, o4-mini at times failed to infer SDoH subcategories that were implied but not explicitly stated; 2) Over-interpretation of insufficient evidence. Conversely, the model also exhibited over-reasoning, assigning labels when the evidence was directionally relevant but falls short of what the annotation guidelines require; 3) Prompt instruction non-adherence. In several cases, the model failed to follow explicit SDoH definitions provided in the prompt; 4) Temporality misclassification. The model occasionally correctly identified an SDoH mention but failed to determine whether it reflects the patient’s current or historical status. The first two error types reflect a gap between the model and human annotators regarding what constitutes sufficient evidence for SDoH classification, suggesting the model lacks a calibrated threshold aligned with the annotation framework. Future efforts should focus on closing this gap through refined SDoH definitions and boundary-clarifying examples.

## DISCUSSION

### Principal Findings

This study examined the extraction of SDoH information across seven domain categories, including less studied domains such as social resources and health insurance status, and 23 corresponding subcategories from discharge summaries and progress notes in a large health system. We evaluated seven GPT models using under multiple prompting strategies and compared their performance with an expert-designed, iteratively-optimized rule-based system (RBS). Recently released GPT models with improved reasoning capabilities, such as GPT-5 and o4-mini, achieved the best overall performance at both the domain-category and subcategory levels. While RBS demonstrated high precision for domain categories, it exhibited consistently low recall due to the inherent rigidity of manually engineered rules. For example, GPT models correctly identified “patient needs WIC” (WIC: Women, Infants, & Children Nutrition Program) as an indicator of “Food insecurity” by recognizing WIC’s role as a food assistance program, while appropriately excluding gastrointestinal-related eating difficulties as medical rather than socioeconomic issues. In contrast, RBS failed to capture the WIC reference due to its reliance on predefined lexicons and misclassified gastrointestinal-related eating problems as food insecurity, lacking the semantic understanding to distinguish medical from social determinants. Our findings differ from Patra et al. [19], who reported superior RBS performance over LLM-based approach for classifying social support and social isolation, primarily because their RBS closely mirrored the gold-standard annotation rule book, leading to overfitting.

Focusing on our GPT models, the domain-level macro-F1 scores of 0.82-0.89 are broadly consistent with recent evaluations of LLM-based SDoH extraction [15,17,20]. For example, Keloth et al. [17] reported macro-F1 scores ranging from 0.53 to 0.84 across four institutions using instruction fine-tuned LLaMA models, while Guevara et al. [15] achieved a macro-F1 of 0.71 for sentence-level SDoH classification using fine-tuned Flan-T5 models. At the subcategory level, our GPT models achieved macro-F1 scores of 0.78-0.88 across 23 subcategories. In contrast, Keloth et al. reported lower Level-2 macro-F1 (0.45-0.59), though their task additionally required temporality determination for some categories. Earlier-generation LLMs have shown more limited in-context learning performance, with GPT-4 one-shot prompting achieving only 0.65 micro-averaged F1 on the n2c2/UW SHAC event extraction task [25,55] and few-shot prompted LLaMA-2 underperforming fine-tuned models [17]. While these comparisons should be interpreted with caution due to differences in SDoH taxonomies, annotation schemas, note types, evaluation metrics, and the enriched nature of our validation set, the overall pattern suggests that recent reasoning-capable GPT models, when applied to pre-screened clinical text in prompted settings, can achieve performance comparable to that reported for fine-tuned approaches and represent a meaningful advance over earlier LLM prompting-based models. Additional evaluation on 500 unfiltered text segments, drawn from notes with a median documentation year of 2014 (IQR 2010-2018), showed low false-positive rates and provided preliminary evidence that the model can identify SDoH concepts in unfiltered clinical text, though the small number of SDoH-positive cases (n=42) precludes definitive conclusions about broader generalizability (Appendix S7).

Comparing efficiency in model development, GPT-based approaches in zero-shot or few-shot settings required substantially less development time and cost than constructing the RBS, which involved iterative rule creation and refinement. Among the GPT models, newer “mini” models, such as GPT-5-mini and o4-mini, which retain reasoning capabilities, performed competitively with OpenAI’s flagship GPT-5 model while demonstrating substantially lower inference costs and latency. In our benchmarking (Appendix S5), o4-mini (5-shot) and GPT-5-mini (5-shot) incurred estimated costs of $0.008 and $0.005 per text segment, respectively, compared with $0.029 for GPT-5 (5-shot), representing 72-82% cost reductions. In contrast, earlier small models such as GPT-4o-mini and GPT-4.1-mini, though offer even lower per-token pricing, did not perform on-par with their respective full models. At the observed single-worker throughput for o4-mini (5-shot), processing 100,000 and 1,000,000 segments would require approximately 10 days and 102 days, respectively. In practice, cloud providers offer asynchronous batch processing with higher rate-limit pools and a 50% cost discount, which could substantially reduce wall-clock time relative to sequential single-worker deployment. However, realized throughput remains subject to API quotas, particularly for reasoning models such as o4-mini, whose hidden reasoning tokens increase token consumption. Alternatively, LLM-generated annotations on a representative subset could be used to train a local classifier, reducing API dependence for large-scale deployment.

Our prompt engineering evaluation highlighted the sensitivity of model performance to the wording of prompts, showing a precision-recall trade-off tunable through stricter or more liberal prompt styles. For few-shot prompting, when comparing the addition of different types of examples, we found that including cases where annotators required clarification or adjudication, along with explanations, yielded optimal performance for SDoH subcategories, suggesting the value of exposing LLMs to examples that humans themselves found challenging and ambiguous. For example, to improve GPT models’ ability to parse questionnaire text, we provided examples demonstrating how to identify question-answer pairs and when to recognize missing answers, in which case no subcategory should be assigned. Taken together, these findings suggest that adopting a systematic approach of prompt design and incorporating challenging examples should be considered as best practices for LLM-based extraction methods.

Lastly, we evaluated various model ensemble strategies using late fusion. Combining RBS with a committee of GPT models employing different prompting styles improved domain-category extraction compared with using GPT models alone. For example, the RBS-GPT hybrid correctly recognized “involvement in spiritual community” as a mention of “Social resource”, whereas the GPT models alone failed to do so. For subcategories, the GPT-committee demonstrated that precision or recall could be selectively improved depending on the fusion function applied, though without overall performance gain. These findings suggest the complementary strengths of rule-based and LLM-based systems, though the contribution of RBS to the ensemble for domain-category extraction may be inflated by the keyword-enriched validation set. The lack of improvement at the subcategory level also indicates the need for more advanced and adaptive ensemble strategies, such as mixture-of-experts approaches where different models (“experts”) handle different inputs, a direction for future investigation.

### Limitations and Future Directions

This study has several limitations. First, both our RBS and GPT-based models were validated in a single healthcare system, and performance may vary elsewhere depending on documentation practices. This concern may be particularly relevant for the RBS, whose lexicon was filtered and refined through manual review of MGB notes and may therefore be less generalizable to other health systems. Additionally, because the validation set was sampled from the same source corpus, the reported RBS performance may represent an optimistic upper bound in this setting. At the same time, MGB comprises more than eight hospitals and multiple community health centers with heterogeneous catchment areas and clinical practices, supporting some degree of generalizability. Second, our primary goal was to develop a cost-efficient SDoH extraction pipeline for resource-constrained settings, which motivated our focus on rule-based and LLM-prompting based approaches that can be deployed with minimal annotation effort and computational resources. We did not explore the use of synthetic data generation for training supervised models or LLM fine-tuning, which, while potentially reducing annotation cost, still require computation for model training. Future work could examine how these approaches compare. Third, although recent GPT models are considered state-of-the-art for many tasks, particularly those requiring reasoning capabilities, we did not conduct a systematic comparison with open-weight LLMs. Such evaluation would provide a more complete performance assessment. Fourth, we annotated text segments rather than entire clinical notes, as manual review suggested that segments contained sufficient information for SDoH extraction. However, this approach may miss SDoH mentions that depend on long-distance context within full notes. Finally, since we applied rule-based filtering during data sampling, the recall and F1 scores reported from our primary validation set are based on enriched text and therefore do not fully reflect performance in the general population of clinical notes. Although an additional evaluation on 500 unfiltered text segments (Appendix S7) suggested that model performance generalizes beyond the enriched setting, the limited number of SDoH-positive cases in unfiltered text prevents definitive conclusions about generalizability and warrants further validation on larger, independently sampled datasets.

The ability to accurately and efficiently extract SDoH factors from clinical notes has important implications for healthcare systems, as such information is often under-documented in structured EHR data. Our approach provides a cost-efficient method for SDoH extraction, demonstrating consistently high precision and recall across both domain categories and subcategories. By pre-screening clinical notes with a rule-based NLP filter to identify relevant text segments before applying LLM-based extraction, we improved efficiency and substantially reduced computational costs compared with processing entire patient notes directly with LLMs. The approach presented in this study has the potential to advance important population health research and ultimately inform clinical outcomes and evidence-based policy intervention [6,56]. Incorporating SDoH information into clinical prediction models is expected to improve prediction performance, as recent studies suggest [57,58], while also supporting audits of algorithmic biases beyond traditional demographic variables [59]. Finally, as interest grows in implementing such NLP systems in clinical operations, it will be critical to anticipate and mitigate potential unintended consequences of automated SDoH extraction for patients [60].

Future work should assess the robustness and generalizability of our approach to other healthcare systems, especially those with very different clinical settings, patient demographics and socioeconomic environments. Furthermore, social determinants are highly context dependent. Existing models often categorize a patient’s social circumstance into predefined labels without capturing context, limiting their ability to provide a comprehensive and contextually relevant understanding of patient’s social needs. Lybarger et al. [25] employed an event-based annotation scheme characterizing each SDoH mention with multiple arguments (e.g., *employment status, duration, history, and type* for “employment”) from the social history sections. We plan to explore methods for incorporating richer contextual information, moving beyond the contextual attributes used in our rule-based approach and related work [5,17,25]. We will investigate text summarization methods to generate structured representations that encapsulate a comprehensive profile of patient’s SDoH status for each clinical encounter. Lastly, it will also be important to validate the utility of the extracted SDoH information, along with community-level socioeconomic factors [56], in supporting downstream health research applications including suicide prediction [9,58,61] and model auditing [59,62].

## Conclusions

This study presented and compared two NLP approaches, an RBS and GPT-based models, for extracting SDoH information from clinical text segments retrieved from clinical notes. Recent GPT models with advanced reasoning capabilities achieved superior performance in identifying seven SDoH domain categories and 23 subcategories with high precision and recall, requiring no additional training or fine-tuning. In addition, we developed and validated late-fusion ensembles combining both approaches to optimize extraction performance. By making our code and prompts available to the scientific community, we provide a cost-efficient solution for accurate SDoH extraction, with the potential for advancing important downstream health research applications.

## Data Availability

Protected Health Information restrictions apply to the availability of the clinical data here, which were used under IRB approval for use only in the current study. As a result, the data used in the present study is not publicly available.

## ACKNOWLEDGEMENTS

We declare the use of the generative AI tool ChatGPT (OpenAI) for language-editing and formatting of the manuscript.

## DATA AVAILABILITY

Protected Health Information restrictions apply to the availability of the clinical data here, which were used under IRB approval for use only in the current study. As a result, this dataset is not publicly available. The code of our rule-based system and LLM prompts is available at https://github.com/bwang482/SDoH_Extraction.

## AUTHORS’ CONTRIBUTIONS

Conceptualization: B.W., K.W.C., and J.W.S.

Data curation: B.W., D.K.

Formal analysis: B.W.

Funding acquisition: B.W., J.W.S.

Investigation: B.W., K.W.C., and J.W.S.

Methodology: B.W., K.W.C., J.W.S., D.K., and C.R.C.,

Project administration: B.W.

Resources: K.W.C., J.W.S.

Supervision: K.W.C., J.W.S.

Validation: B.W., D.K.

Visualization: B.W.

Writing – original draft: B.W.

Writing – review & editing: B.W., D.K., C.R.C., K.W.C., and J.W.S.

## ABBREVIATIONS

SDoH: social determinants of health
EHR: electronic health record
NLP: natural language processing
LLM: large language model
RBS: rule-based system
GPT: generative pre-trained transformer
MGB: Mass General Brigham
RPDR: Research Patient Data Registry
ICD: International Classification of Diseases

