## Supplementary Materials for "Extracting Social Determinants of Health from Electronic Health Records: Development and Comparison of Rule-Based and Large Language Model Methods"

### Supplementary Material

#### APPENDIX S1: DEVELOPING LEXICON AND RULES

##### S1.1 Surveys and screening tools

- Children's HealthWatch Hunger Vital Sign [1]
- USDA Household Food Security Survey
- The Centers for Medicare & Medicaid Services (CMS) Accountable Health Communities Health-Related Social Needs Screening Tool
- Health Begins Upstream Risks Screening Tool
- RAND MOS Social Support Survey Instrument [2]
- PRAPARE: Protocol for Responding to and Assessing Patient Assets, Risks, and Experiences
- International physical activity questionnaire: 12-country reliability and validity [3]
- The Multi-Ethnic Study of Atherosclerosis (MESA) - Employment Questionnaire [4]
- UCLA Loneliness Scale (revised) [5]
- The Common Fund's Patient-Reported Outcomes Measurement Information System (PROMIS) [6]
- The Health Leads Social Needs Screening Toolkit

##### S1.2 Lexicon filtering and refinement

The initial seed terms were reviewed and filtered by a clinical psychologist specializing in socio-environmental factors of mental health conditions. The expanded lexicon underwent four iterative rounds of manual review:

- Rounds 1-3: Applied pattern matching using our SDoH lexicon and context rules to progressively larger samples (20, 100, 1,000 patients) and reviewed all returned matches.
- Round 4: Applied pattern matching to 4,000 randomly selected patients, then:
    - Ranked query terms in the SDoH lexicon by matching frequency.
    - Reviewed 10 random sampled matches from each of the top 20 terms.
    - Reviewed 10 random sampled matches from 20 randomly selected remaining terms per subcategory.
    - Total: 4,800 reviewed matches.

Before each review round, we removed duplicate or highly similar matched entities and their corresponding sentences using Levenshtein distance. Following all reviews, only query terms achieving greater than 80% positive predictive value (precision) were retained in the final lexicon. The 2,000-patient gold-standard cohort (from which the 226 validation segments were drawn) was held out from any lexicon development to prevent data contamination.

##### S1.3 Components of the Rule-based System

Preprocessor: We implemented a preprocessing module to clean clinical text, particularly for poorly structured questionnaires embedded within notes.

Section detector: We adapted existing section detection patterns to identify sections within clinical notes.

Sentence splitter: We chose pySBD (Python Sentence Boundary Disambiguation; https://github.com/nipunsadvilkar/pySBD) [7] for detecting sentence boundaries in the clinical notes after comparing it with PyRuSH (Rule-based sentence Segmentation using Hashing; https://github.com/medspacy/PyRuSH) [8].

Entity matcher: We utilized medspaCy's TargetMatcher module. (https://github.com/medspacy/target_matcher), which extends spaCy's core pattern matching functionality (https://spacy.io/api/matcher), combined with our comprehensive SDoH lexicon to identify relevant entities in clinical notes.

Context analyzer: We applied the ConText algorithm [9,10] to determine five contextual attributes for each matched entity: negation, experiencer, temporality, hypothetical status, and certainty.

Postprocessor: Finally, we applied postprocessing rules to all matched entities. For example, the rule-based system was configured to ignore matches found in sections unlikely to contain any patient-reported SDoH information, such as instructions and care plan sections.

#### APPENDIX S2: ANNOTATION GUIDELINES

Annotation guidelines:

https://github.com/bwang482/SDoH_Extraction/tree/main/annotation_guideline

#### Appendix S3: LLM prompts

Zero-shot prompt (strict style):

https://github.com/bwang482/SDoH_Extraction/blob/main/prompts_strict.py

Zero-shot prompt (balanced style):

https://github.com/bwang482/SDoH_Extraction/blob/main/prompts_balanced.py

Zero-shot prompt (liberal style):

https://github.com/bwang482/SDoH_Extraction/blob/main/prompts_liberal.py

Few-shot prompt (template only):

https://github.com/bwang482/SDoH_Extraction/blob/main/few-shot_prompt.py

#### APPENDIX S4: GPT MODEL VERSIONS

We list the names and version identifiers of the GPT models evaluated in this study:

| Model name | Model version |
|---|---|
| GPT-4o | 2024-11-20 |
| GPT-4.1 | 2025-04-14 |
| GPT-4.1-mini | 2025-04-14 |
| GPT-5 | 2025-08-07 |
| GPT-5-mini | 2025-08-07 |
| GPT-o3 | 2025-04-16 |
| GPT-o4-mini | 2025-04-16 |

#### APPENDIX S5: GPT MODEL INFERENCE TIME AND COST COMPARISON

Token usage reported below represents the mean number of tokens per text segment. Specifically, input (prompt) tokens, API-reported reasoning tokens, and visible output tokens are summarized as averages across all segments. Inference time and estimated inference cost are similarly reported as mean values per segment. Completion tokens correspond to the sum of API-reported reasoning tokens and visible output tokens. Estimated inference cost was calculated as

$$(input\ tokens \times input\ token\ rate) + (completion\ tokens \times output\ token\ rate)$$

based on Azure OpenAI pricing. All experiments were conducted using an Azure OpenAI deployment in the East US region with the reasoning effort set to high and a single-worker, sequential inference setup.

Azure OpenAI token pricing used during study period:

| Model | Input (per 1M tokens) | Output (per 1M tokens) |
|---|---|---|
| O4-mini | $1.10 | $4.40 |
| GPT-5-mini | $0.25 | $2.00 |
| GPT-5 | $1.25 | $10.00 |

Mean token usage, inference time, and estimated cost per text segment by model and prompting strategy:

| Model | Prompting strategy | Input tokens | Reasoning tokens | Output tokens | Inference time (s) | Est. inference cost ($) |
|---|---|---|---|---|---|---|
| O4-mini | Zero-shot | 2441.54 | 766.02 | 45.45 | 6.98 | 0.006 |

| | | | | | | |
|---|---|---|---|---|---|---|
| GPT-5-mini | Zero-shot | 2441.54 | 1438.02 | 44.17 | 14.02 | 0.004 |
| GPT-5 | Zero-shot | 2441.54 | 1997.03 | 42.31 | 32.98 | 0.023 |
| O4-mini | 5-shot | 3392.54 | 986.62 | 45.69 | 8.89 | 0.008 |
| GPT-5-mini | 5-shot | 3392.54 | 1815.22 | 44.50 | 22.05 | 0.005 |
| GPT-5 | 5-shot | 3392.54 | 2462.02 | 42.61 | 34.64 | 0.029 |

#### APPENDIX S6: PERFORMANCE ACROSS DEMOGRAPHIC GROUPS

When stratified by gender, self-reported race, and ethnicity, permutation tests did not detect statistically significant differences in performance across demographic groups for o4-mini in identifying SDoH domain categories and subcategories (Table S6-S8). However, several subgroups were small (e.g., n=17 for Hispanic patients), substantially limiting statistical power and increasing the risk of Type II error. These results should therefore not be interpreted as evidence that the model is free of demographic bias, and a larger, more demographically balanced dataset is needed for robust bias evaluation.

#### APPENDIX S7: PERFORMANCE ON UNFILTERED CLINICAL TEXT

To assess LLM performance on unfiltered clinical text, we randomly sampled 500 text segments from the stratified 2,000-patient cohort without applying keyword pre-screening. Of these, 42 segments (8.4%) contained at least one SDoH domain category, and 34 (6.8%) contained at least one subcategory, confirming the expected sparsity of SDoH mentions in general clinical text. On this "control set", o4-mini (5-shot), the best performing model, achieved macro-averaged precision/recall/F1 of 0.94/0.75/0.83 for domain categories and 0.83/0.87/0.85 for subcategories. Additionally, we evaluated its performance on the segments containing no SDoH information ("negative control set"). The model incorrectly identified only 3 of 458 and 4 of 466 negative segments at the domain-category and subcategory levels, respectively (FPR = 0.01 for both), indicating minimal hallucination on these negative control samples. These results on unfiltered clinical text are promising, particularly the low false positive rate on negative segments, but because the number of SDoH-positive segments was small, they should be considered preliminary rather than definitive evidence of broader generalizability.

O4-mini (5-shot) performance on randomly sampled, unfiltered clinical text segments (N = 500). FP: false positive; FPR: false positive rate.

| SDoH domain categories | | | | | SDoH subcategories | | | | |
|---|---|---|---|---|---|---|---|---|---|
| P | R | F1 | FP count | FPR | P | R | F1 | FP count | FPR |
| 0.94 | 0.75 | 0.83 | 3 | 0.01 | 0.83 | 0.87 | 0.85 | 4 | 0.01 |

#### Supplementary Tables

Table S1: Overall performance (macro-averaged) comparison between rule-based and GPT-based models.

Table S2: Overall performance (micro-averaged) comparison between rule-based and GPT-based models.

Table S3: F1 scores across SDoH domain categories and subcategories.

Table S4: Performance across different prompt styles (strict, balanced, and liberal).

Table S5: Performance across few-shot prompting strategies with different example types.

Table S6: F1 scores of o4-mini (5-shot) across SDoH domains by demographic group.

Table S7: F1 scores of o4-mini (5-shot) across SDoH categories by self-reported race.

Table S8: F1 scores of o4-mini (5-shot) across SDoH subcategories by demographic group.

Table S9: F1 scores of our rule-based system across SDoH categories by demographic group.

Table S10: F1 scores of our rule-based system across SDoH categories by self-reported race.

Table S11: F1 scores of rule-based system across SDoH subcategories by demographic group.

Table S12: ICD Z and V codes related to SDoH.

Table S13: Selected ICD Z and V codes specifically relevant to each SDoH category.

Table S14: Primary error types of RBS and o4-mini (5-shot).

Table S15: Micro-averaged performance and absolute error counts across fusion strategies.

**Table S1.** Overall performance comparison between rule-based and GPT-based approaches using macro-averaged precision, recall, and F1 scores.

Note: Unless otherwise indicated, reasoning models were run with the high-reasoning setting. The o4-mini low-reasoning run is labeled explicitly for comparison.

| Models | SDoH domain categories | | | SDoH subcategories | | |
|---|---|---|---|---|---|---|
| | Precision | Recall | F1 | Precision | Recall | F1 |
| RBS | 0.96 | 0.68 | 0.78 | 0.69 | 0.58 | 0.59 |
| GPT-4o (zero-shot) | 0.95 | 0.80 | 0.87 | 0.77 | 0.87 | 0.81 |
| GPT-4o (5-shot) | 0.95 | 0.80 | 0.86 | 0.83 | 0.87 | 0.84 |
| GPT-4.1 (zero-shot) | 0.93 | 0.80 | 0.86 | 0.78 | 0.87 | 0.82 |
| GPT-4.1 (5-shot) | 0.94 | 0.78 | 0.85 | 0.84 | 0.85 | 0.83 |
| GPT-4.1-mini (zero-shot) | 0.91 | 0.84 | 0.86 | 0.73 | 0.88 | 0.78 |
| GPT-4.1-mini (5-shot) | 0.94 | 0.81 | 0.87 | 0.79 | 0.85 | 0.80 |
| o4-mini (low reasoning, zero-shot) | 0.93 | 0.75 | 0.83 | 0.80 | 0.87 | 0.83 |
| o4-mini (zero-shot) | 0.95 | 0.75 | 0.84 | 0.85 | 0.91 | 0.87 |
| o4-mini (5-shot) | 0.98 | 0.76 | 0.86 | 0.90 | 0.87 | 0.88 |
| o3 (zero-shot) | 0.96 | 0.74 | 0.83 | 0.87 | 0.90 | 0.87 |
| o3 (5-shot) | 0.95 | 0.74 | 0.83 | 0.86 | 0.87 | 0.86 |
| GPT-5-mini (zero-shot) | 0.91 | 0.81 | 0.85 | 0.80 | 0.92 | 0.84 |
| GPT-5-mini (5-shot) | 0.91 | 0.87 | 0.89 | 0.82 | 0.88 | 0.84 |

| | | | | | | |
|---|---|---|---|---|---|---|
| GPT-5 (zero-shot) | 0.95 | 0.72 | 0.82 | 0.86 | 0.89 | 0.87 |
| GPT-5 (5-shot) | 0.96 | 0.84 | 0.89 | 0.90 | 0.85 | 0.86 |

**Table S2.** Overall performance comparison between rule-based and GPT-based approaches using micro-averaged precision, recall, and F1 scores.

Note: Unless otherwise indicated, reasoning models were run with the high-reasoning setting. The o4-mini low-reasoning run is labeled explicitly for comparison.

| Models | SDoH domain categories | | | SDoH subcategories | | |
|---|---|---|---|---|---|---|
| | Precision | Recall | F1 | Precision | Recall | F1 |
| Rule-based (RBS) | 0.97 | 0.63 | 0.76 | 0.67 | 0.49 | 0.57 |
| GPT-4o (zero-shot) | 0.96 | 0.81 | 0.88 | 0.77 | 0.85 | 0.81 |
| GPT-4o (5-shot) | 0.96 | 0.81 | 0.88 | 0.82 | 0.84 | 0.83 |
| GPT-4.1-mini (zero-shot) | 0.92 | 0.84 | 0.88 | 0.73 | 0.84 | 0.78 |
| GPT-4.1-mini (5-shot) | 0.95 | 0.82 | 0.88 | 0.78 | 0.81 | 0.80 |
| GPT-4.1 (zero-shot) | 0.95 | 0.81 | 0.87 | 0.79 | 0.85 | 0.82 |
| GPT-4.1 (5-shot) | 0.96 | 0.77 | 0.86 | 0.84 | 0.83 | 0.83 |
| o4-mini (low reasoning, zero-shot) | 0.95 | 0.77 | 0.85 | 0.79 | 0.87 | 0.83 |
| o4-mini (zero-shot) | 0.96 | 0.76 | 0.85 | 0.84 | 0.90 | 0.87 |
| o4-mini (5-shot) | 0.98 | 0.76 | 0.86 | 0.88 | 0.88 | 0.88 |
| o3 (zero-shot) | 0.97 | 0.76 | 0.85 | 0.84 | 0.89 | 0.86 |
| o3 (5-shot) | 0.97 | 0.76 | 0.85 | 0.84 | 0.87 | 0.85 |
| GPT-5-mini (zero-shot) | 0.92 | 0.82 | 0.87 | 0.75 | 0.93 | 0.83 |
| GPT-5-mini (5-shot) | 0.92 | 0.86 | 0.89 | 0.79 | 0.90 | 0.84 |
| GPT-5 (zero-shot) | 0.96 | 0.74 | 0.84 | 0.84 | 0.88 | 0.86 |
| GPT-5 (5-shot) | 0.97 | 0.83 | 0.89 | 0.88 | 0.84 | 0.86 |

**Table S3.** F1 scores across SDoH domain categories and subcategories.

Models include our rule-based system, GPT-4o (zero-shot/0s), GPT-4o (five-shot/5s), GPT-4.1 (zero-shot/0s), GPT-4.1 (five-shot/5s), o4-mini (zero-shot/0s), and o4-mini (five-shot/5s).

| SDoH | RBS | 4o (0s) | 4o (5s) | 4.1 (0s) | 4.1 (5s) | o4-mini (0s) | o4-mini (5s) |
|---|---|---|---|---|---|---|---|
| Social resources | 0.61 | 0.85 | 0.84 | 0.87 | 0.82 | 0.83 | 0.86 |
| - *Good social resources* | 0.33 | 0.76 | 0.78 | 0.77 | 0.75 | 0.84 | 0.86 |
| - *Poor social resources* | 0.56 | 0.67 | 0.58 | 0.67 | 0.67 | 0.67 | 0.70 |
| - *Living with or accompanied by someone* | 0.31 | 0.82 | 0.85 | 0.87 | 0.90 | 0.93 | 0.90 |
| - *Living alone* | 0.96 | 0.97 | 0.97 | 0.97 | 0.97 | 0.97 | 0.97 |
| Physical activity | 0.74 | 0.94 | 0.91 | 0.88 | 0.90 | 0.85 | 0.89 |
| - *Physically active* | 0.73 | 0.89 | 0.86 | 0.83 | 0.80 | 0.92 | 0.88 |
| - *Not/barely physically active* | 0.64 | 0.93 | 0.97 | 0.93 | 0.90 | 0.93 | 0.97 |
| General financial status | 0.81 | 0.78 | 0.78 | 0.75 | 0.76 | 0.73 | 0.85 |
| - *General financial security* | 0.54 | 0.76 | 0.73 | 0.76 | 0.76 | 0.80 | 0.86 |
| - *Financial insecurity* | 0.50 | 0.92 | 0.91 | 0.78 | 0.80 | 0.74 | 0.91 |
| Employment status | 0.75 | 0.92 | 0.93 | 0.92 | 0.89 | 0.90 | 0.87 |
| - *Stable employment* | 0.46 | 0.84 | 0.86 | 0.86 | 0.90 | 0.94 | 0.91 |

| | | | | | | | |
|---|---|---|---|---|---|---|---|
| *- Unemployment* | 0.74 | 0.74 | 0.81 | 0.82 | 0.85 | 0.77 | 0.79 |
| *- Job loss* | 0.89 | 0.88 | 0.94 | 0.89 | 0.88 | 0.94 | 0.80 |
| *- Disability and inability to work* | 0.63 | 0.83 | 0.87 | 0.83 | 0.83 | 0.87 | 0.86 |
| *- Retirement* | 0.85 | 0.88 | 0.92 | 0.92 | 0.96 | 1.0 | 1.0 |
| *- Other job insecurity and employment issues* | 0.50 | 0.78 | 0.74 | 0.88 | 0.71 | 0.88 | 0.80 |
| Housing status | 0.92 | 0.94 | 0.94 | 0.88 | 0.92 | 0.91 | 0.83 |
| *- Secure/quality housing* | 0.50 | 0.82 | 0.90 | 0.78 | 0.90 | 0.86 | 0.95 |
| *- Homeless or transitional housing* | 0.70 | 0.67 | 0.88 | 0.67 | 0.88 | 0.82 | 0.86 |
| *- Subsidized/Public Housing* | 0.50 | 0.86 | 0.86 | 0.77 | 0.77 | 0.92 | 0.91 |
| *- Other housing instability issues* | 0.54 | 0.69 | 0.77 | 0.67 | 0.71 | 0.76 | 0.83 |
| Food security status | 0.80 | 0.76 | 0.76 | 0.78 | 0.76 | 0.80 | 0.80 |
| *- Food security* | 0.12 | 0.77 | 0.77 | 0.77 | 0.83 | 0.92 | 0.92 |
| *- Food insecurity* | 0.33 | 0.78 | 0.86 | 0.75 | 0.80 | 0.87 | 0.90 |
| Insurance status | 0.87 | 0.89 | 0.90 | 0.91 | 0.89 | 0.85 | 0.89 |
| *- General insurance coverage* | 0.82 | 0.80 | 0.87 | 0.88 | 0.88 | 0.88 | 0.91 |
| *- Lack of insurance* | 0.71 | 0.73 | 0.76 | 0.76 | 0.84 | 0.84 | 0.84 |
| *- Government-assisted insurance* | 0.67 | 0.86 | 0.83 | 0.92 | 0.80 | 1.0 | 1.0 |

**Table S4**. Performance across different prompt styles (strict, balanced, and liberal) using macro-averaged precision, recall, and F1 scores.

| Prompt for o4-mini | SDoH domain categories | | | SDoH subcategories | | |
|---|---|---|---|---|---|---|
| | Precision | Recall | F1 | Precision | Recall | F1 |
| Strict (zero-shot) | 0.95 | 0.75 | 0.84 | 0.85 | 0.91 | 0.87 |
| Balanced (zero -shot) | 0.95 | 0.81 | 0.87 | 0.79 | 0.90 | 0.83 |
| Liberal (zero -shot) | 0.87 | 0.88 | 0.88 | 0.69 | 0.95 | 0.79 |
| Strict (5-shot) | 0.98 | 0.76 | 0.86 | 0.90 | 0.87 | 0.88 |
| Balanced (5 -shot) | 0.95 | 0.81 | 0.87 | 0.81 | 0.89 | 0.84 |
| Liberal (5 -shot) | 0.92 | 0.86 | 0.88 | 0.78 | 0.93 | 0.84 |

**Table S5**. Performance across few-shot prompting strategies with different example types using macro-averaged precision, recall, and F1 scores.

Each of the easy and hard example sets contains 5 examples.

| Strategy | SDoH domain categories | | | SDoH subcategories | | |
|---|---|---|---|---|---|---|
| | Precision | Recall | F1 | Precision | Recall | F1 |
| Easy examples | 0.95 | 0.74 | 0.83 | 0.86 | 0.89 | 0.87 |
| Easy examples with explanations | 0.96 | 0.74 | 0.83 | 0.86 | 0.89 | 0.87 |
| Hard examples | 0.97 | 0.72 | 0.82 | 0.88 | 0.87 | 0.87 |
| Hard examples with explanations | 0.98 | 0.76 | 0.86 | 0.90 | 0.87 | 0.88 |
| Synthetic examples | 0.98 | 0.70 | 0.81 | 0.88 | 0.87 | 0.87 |
| Synthetic examples with explanations | 0.96 | 0.72 | 0.82 | 0.88 | 0.90 | 0.88 |

**Table S6.** F1 scores of o4-mini (5-shot) across SDoH domains by demographic group.

M: Male; F: Female; W: Self-reported White; NW: Self-report non-White; H: Hispanic; NH: Non-Hispanic. Statistical significance was assessed using a patient-level cluster permutation test on macro-averaged F1 (10,000 iterations) to account for within-patient correlation. No statistically significant differences in model performance were found between demographic groups (α = 0.05).

| SDoH | Overall | M | F | W | NW | H | NH |
|---|---|---|---|---|---|---|---|
| Social resources | *0.86* | 0.89 | 0.83 | 0.84 | 0.87 | 1.0 | 0.84 |
| Physical activity | *0.89* | 0.76 | 1.0 | 0.94 | 0.86 | 1.0 | 0.88 |
| General financial status | *0.85* | 0.86 | 0.83 | 0.90 | 0.83 | 0.86 | 0.85 |
| Employment status | *0.87* | 0.91 | 0.85 | 0.88 | 0.87 | 0.86 | 0.87 |
| Housing status | *0.83* | 0.88 | 0.78 | 0.82 | 0.84 | 1.0 | 0.81 |
| Food security status | *0.80* | 0.88 | 0.70 | 0.92 | 0.75 | 0.75 | 0.81 |
| Insurance status | *0.89* | 0.83 | 0.94 | 0.92 | 0.88 | 1.0 | 0.89 |
| *Permutation test* | | *P-value = 0.797* | | *P-value = 0.252* | | *P-value = 0.143* | |

**Table S7.** F1 scores of o4-mini (5-shot) across SDoH categories by self-reported race.

The self-reported race groups are: “White”, “Black/African American”, “Asian” and “Other”. Statistical significance was assessed using a patient-level cluster permutation test on macro-averaged F1 (10,000 iterations) to account for within-patient correlation. No statistically significant differences in model performance were found between self-reported race groups ($\alpha = 0.05$).

| SDoH | Overall | White | Black/AA | Asian | Other |
|---|---|---|---|---|---|
| Social resources | *0.86* | 0.84 | 0.74 | 0.90 | 0.94 |
| Physical activity | *0.89* | 0.94 | 0.82 | 0.86 | 0.90 |
| General financial status | *0.85* | 0.90 | 0.80 | 0.83 | 0.91 |
| Employment status | *0.87* | 0.88 | 0.90 | 0.81 | 0.89 |
| Housing status | *0.83* | 0.82 | 0.81 | 0.86 | 0.91 |
| Food security status | *0.80* | 0.92 | 0.92 | 0.67 | 0.60 |
| Insurance status | *0.89* | 0.92 | 0.84 | 0.94 | 0.88 |
| *Permutation test vs self-reported White group* | | | *P-value = 0.285* | *P-value = 0.394* | *P-value = 0.60* |

**Table S8.** F1 scores of o4-mini (5-shot) across SDoH subcategories by demographic group.

M: Male; F: Female; W: Self-reported White; NW: Self-report non-White. Statistical significance was assessed using a patient-level cluster permutation test on macro-averaged F1 (10,000 iterations) to account for within-patient correlation. No statistically significant differences in model performance were found between demographic groups (α = 0.05). N/A indicates zero instances of this subcategory in the demographic subgroup.

| SDoH | Sub-categories | Overall | M | F | W | NW |
|---|---|---|---|---|---|---|
| Social resources | Good social resources | 0.86 | 0.94 | 0.73 | 0.78 | 0.89 |
| | Poor social resources | 0.70 | 0.50 | 0.91 | 0.89 | 0.57 |
| | Living with or accompanied by someone | 0.90 | 0.91 | 0.89 | 0.80 | 0.93 |
| | Living alone | 0.97 | 1.0 | 0.96 | 1.0 | 0.96 |
| Physical activity | Physically active | 0.88 | 0.93 | 0.80 | 0.80 | 0.90 |
| | Not/barely physically active | 0.97 | 1.0 | 0.95 | 0.91 | 1.0 |
| General financial status | General financial security | 0.86 | 0.94 | 0.80 | 1.0 | 0.81 |
| | Financial insecurity | 0.91 | 0.83 | 1.0 | 1.0 | 0.88 |

| | | | | | | |
|---|---|---|---|---|---|---|
| Employment status | Stable employment | 0.91 | 0.91 | 0.90 | 0.88 | 0.92 |
| | Unemployment | 0.79 | 0.80 | 0.78 | 0.80 | 0.78 |
| | Job loss | 0.80 | 1.0 | 0.73 | N/A | 0.80 |
| | Disability and inability to work | 0.86 | 0.80 | 0.88 | 0.50 | 0.94 |
| | Retirement | 1.0 | 1.0 | 1.0 | 1.0 | 1.0 |
| | Other job insecurity and employment issues | 0.80 | 0.83 | 0.67 | 1.0 | 0.67 |
| Housing status | Secure/quality housing | 0.95 | 0.93 | 1.0 | 1.0 | 0.91 |
| | Homeless/transitional housing | 0.86 | 0.91 | 0.67 | 0.80 | 1.0 |
| | Subsidized/Public Housing | 0.91 | 1.0 | 0.89 | 1.0 | 0.80 |
| | Other housing instability issues | 0.83 | 0.83 | 0.82 | 0.57 | 0.91 |
| Food security status | Food security | 0.92 | 0.91 | 1.0 | 1.0 | 0.89 |
| | Food insecurity | 0.90 | 1.0 | 0.86 | 1.0 | 0.86 |

| | | | | | | |
|---|---|---|---|---|---|---|
| Insurance status | General insurance coverage | 0.91 | 0.92 | 0.89 | 1.0 | 0.89 |
| | Lack of insurance | 0.84 | 0.67 | 0.92 | 1.0 | 0.80 |
| | Government-assisted insurance | 1.0 | 1.0 | 1.0 | 1.0 | 1.0 |
| *Permutation test* | | | *P-value = 0.582* | | *P-value = 0.446* | |

**Table S9.** F1 scores of our rule-based system across SDoH categories by demographic group.

M: Male; F: Female; W: Self-reported White; NW: Self-report non-White; H: Hispanic; NH: Non-Hispanic. Statistical significance was assessed using a patient-level cluster permutation test on macro-averaged F1 (10,000 iterations) to account for within-patient correlation. No statistically significant differences in model performance were found between demographic groups (α = 0.05).

| SDoH | Overall | M | F | W | NW | H | NH |
|---|---|---|---|---|---|---|---|
| Social resources | *0.61* | 0.66 | 0.56 | 0.55 | 0.63 | 0.59 | 0.61 |
| Physical activity | *0.74* | 0.87 | 0.58 | 0.88 | 0.68 | 0.67 | 0.75 |
| General financial status | *0.81* | 0.76 | 0.85 | 0.78 | 0.82 | 0.86 | 0.80 |
| Employment status | *0.75* | 0.70 | 0.78 | 0.74 | 0.75 | 0.77 | 0.75 |
| Housing status | *0.92* | 0.92 | 0.91 | 0.90 | 0.93 | 1.0 | 0.90 |
| Food security status | *0.80* | 0.96 | 0.61 | 1.0 | 0.72 | 0.67 | 0.83 |
| Insurance status | *0.87* | 0.87 | 0.86 | 0.86 | 0.87 | 1.0 | 0.86 |
| *Permutation test* | | *P-value = 0.318* | | *P-value = 0.295* | | *P-value = 0.607* | |

**Table S10.** F1 scores of our rule-based system across SDoH categories by self-reported race.

The self-reported race groups are: “White”, “Black/African American”, “Asian” and “Other”. Statistical significance was assessed using a patient-level cluster permutation test on macro-averaged F1 (10,000 iterations) to account for within-patient correlation. No statistically significant differences in model performance were found between self-reported race groups ($\alpha = 0.05$).

| SDoH | Overall | White | Black/AA | Asian | Other |
|---|---|---|---|---|---|
| Social resources | *0.61* | 0.55 | 0.59 | 0.72 | 0.56 |
| Physical activity | *0.74* | 0.88 | 0.67 | 0.75 | 0.67 |
| General financial status | *0.81* | 0.78 | 0.81 | 0.83 | 0.80 |
| Employment status | *0.75* | 0.74 | 0.65 | 0.78 | 0.81 |
| Housing status | *0.92* | 0.90 | 0.94 | 0.67 | 1.0 |
| Food security status | *0.80* | 1.0 | 0.71 | 0.80 | 0.67 |
| Insurance status | *0.87* | 0.86 | 0.78 | 0.82 | 1.0 |
| *Permutation test vs self-reported White group* | | | *p-value = 0.134* | *p-value = 0.379* | *p-value = 0.605* |

**Table S11.** F1 scores of the rule-based system across SDoH subcategories by demographic group.

M: Male; F: Female; W: Self-reported White; NW: Self-report non-White. Statistical significance was assessed using a patient-level cluster permutation test on macro-averaged F1 (10,000 iterations) to account for within-patient correlation. No statistically significant differences in model performance were found between demographic groups (α = 0.05). N/A indicates zero instances of this subcategory in the demographic subgroup.

| SDoH | Sub-categories | Overall | M | F | W | NW |
|---|---|---|---|---|---|---|
| Social resources | Good social resources | 0.33 | 0.44 | 0.11 | 0.15 | 0.39 |
| | Poor social resources | 0.56 | 0.20 | 0.80 | 0.57 | 0.56 |
| | Living with or accompanied by someone | 0.31 | 0.40 | 0.21 | 0.25 | 0.32 |
| | Living alone | 0.96 | 1.0 | 0.95 | 1.0 | 0.96 |
| Physical activity | Physically active | 0.73 | 0.71 | 0.75 | 0.57 | 0.80 |
| | Not/barely physically active | 0.64 | 0.80 | 0.53 | 0.89 | 0.50 |
| General financial status | General financial security | 0.54 | 0.50 | 0.57 | 0.75 | 0.44 |
| | Financial insecurity | 0.50 | 0.53 | 0.46 | 0.67 | 0.45 |

| | | | | | | |
|---|---|---|---|---|---|---|
| Employment status | Stable employment | 0.46 | 0.43 | 0.48 | 0.44 | 0.46 |
| | Unemployment | 0.74 | 0.75 | 0.74 | 0.80 | 0.71 |
| | Job loss | 0.89 | 1.0 | 0.86 | N/A | 0.89 |
| | Disability and inability to work | 0.63 | 0.40 | 0.71 | 0.0 | 0.75 |
| | Retirement | 0.85 | 0.77 | 0.92 | 0.86 | 0.84 |
| | Other job insecurity and employment issues | 0.50 | 0.44 | 0.67 | 0.67 | 0.33 |
| Housing status | Secure/quality housing | 0.50 | 0.55 | 0.44 | 0.86 | 0.31 |
| | Homeless/transitional housing | 0.70 | 0.80 | 0.40 | 0.77 | 0.57 |
| | Subsidized/Public Housing | 0.50 | 0.40 | 0.55 | 0.67 | 0.40 |
| | Other housing instability issues | 0.54 | 0.73 | 0.40 | 0.25 | 0.67 |
| Food security status | Food security | 0.12 | 0.22 | 0.0 | 0.29 | 0.0 |
| | Food insecurity | 0.33 | 0.31 | 0.36 | 0.0 | 0.42 |

| | | | | | | |
|---|---|---|---|---|---|---|
| Insurance status | General insurance coverage | 0.82 | 0.92 | 0.67 | 0.80 | 0.82 |
| | Lack of insurance | 0.71 | 0.80 | 0.67 | 0.67 | 0.71 |
| | Government-assisted insurance | 0.67 | 0.33 | 0.89 | 0.67 | 0.67 |
| *Permutation test* | | | *P-value = 0.808* | | *P-value = 0.605* | |

**Table S12.** ICD Z and V codes related to SDoH.

| ICD codes | Definition |
|---|---|
| ICD-9: V60* | Housing household and economic circumstances. |
| ICD-9: V61* | Other family circumstances. |
| ICD-9: V62* | Other psychosocial circumstances, e.g., 'Unemployment' or 'Social maladjustment'. |
| ICD-10: Z* | Factors influencing health status and contact with health services. |

**Table S13.** Selected ICD Z and V codes specifically relevant to each SDoH category.

| ICD codes | Definition |
|---|---|
| Social Resources/Connection | |
| Z60.0 | Problems of social environment (used when social circumstances adversely affect the patient). |
| Z60.2 | Problems related to living alone (can be used when isolation or lack of support is identified). |
| Z63.8 | Other specified problems related to primary support groups (a catch-all for issues related to family or social support). |
| V60.3 | Person living alone. |
| V62.4 | Social exclusion or rejection. |
| Physical Activity | |
| Z72.3 | Lack of physical exercise. |
| General Financial Status | |
| Z59.5 | Extreme poverty. |
| Z59.6 | Low income. |
| Employment Status | |
| Z56.0 | Unemployment, unspecified. |
| Z56.1 | Change of job. |
| Z56.2 | Threat of job loss. |
| Z56.3 | Stressful work schedule. |
| Z56.4 | Discord with boss and workmates. |
| Z56.5 | Uncongenial work environment. |
| Z56.6 | Other and unspecified problems related to employment. |
| V62.0 | Unemployment. |
| Housing Status | |

| | |
|---|---|
| Z59.0 | Homelessness. |
| Z59.1 | Inadequate housing. |
| V60.x | Housing related. |
| Food Security Status | |
| Z59.4 | Lack of adequate food and safe drinking water. |
| Insurance Status | |
| Z59.84 | Lack of adequate insurance coverage. |

**Table S14.** Primary error types of the RBS and o4-mini (5-shot) models.

| Approach | Error Type | Direction | Example |
|---|---|---|---|
| RBS | Inability to interpret semantic meaning beyond keywords | FN | Unable to recognize that "patient owns multiple homes" indicates secure housing or infer a patient's current unemployment from a statement describing plans to return to work. |
| | Struggle with abbreviations and domain-specific terms | FP | "Section 8" not recognized as a government-funded rent subsidy program. |
| | Failure to recognize negations | FP | "No exercise intolerance" misidentified as physical inactivity |
| | Failure to recognize temporality | FP | Historical homelessness misidentified as current. |
| | Failure to recognize hypotheticals | FP | "Considering retirement" misidentified as retired. |
| | Failure to distinguish patient from others | FP + FN | Family member's employment attributed to patient. |
| | Semantic disambiguation | FP | "systolic (retired)" misidentified as employment retirement; diet-related mentions misidentified as food insecurity. |
| | Misidentification from question text | FP | Screening question about housing misidentified as patient's housing status, even without a patient response. |
| O4-mini | Missing implicit evidence | FN | Food delivered by family not recognized as food security. |

| | | | |
|---|---|---|---|
| | Over-interpretation of insufficient evidence | FP | Having income treated as sufficient evidence for financial security. |
| | Prompt instruction non-adherence | FP + FN | “Works part-time” classified as stable employment despite prompt explicitly defining it as full-time employment. |
| | Temporality misclassification | FP | Text containing a past date indicating historical lack of insurance, but model classified it as current. |

**Table S15.** Micro-averaged performance metrics and absolute error counts across fusion strategies. Absolute counts denote true positives (TP), false positives (FP), and false negatives (FN) in relative to gold-standard annotations. For domain categories, TP + FN sums to 403 rather than the 410 total annotations because 7 annotations labeled "None of the Above" (NOA) do not contribute to TP or FN for any positive domain category. For subcategories, TP + FN sums to 322 rather than the 475 total annotations because 153 annotations labeled "NA" do not contribute to TP or FN for any positive subcategory label. Analogously, system predictions labeled NOA or NA do not contribute to TP or FP; for example, Majority Vote → Union produced 402 domain-category predictions, of which 21 were NOA, yielding the 381 (TP + FP) reported in the table below.

| GPT-committee → RBS-GPT Fusion | SDoH domain categories | | | | | |
|---|---|---|---|---|---|---|
| | Precision | Recall | F1 | TP | FP | FN |
| Majority voting → ∪ | <u>0.94</u> | <u>0.89</u> | **<u>0.92</u>** | 359 | 22 | 44 |
| Majority voting → ∩ | 0.99 | 0.57 | 0.72 | 228 | 1 | 175 |
| Union → ∪ | 0.90 | 0.93 | 0.91 | 373 | 40 | 30 |
| Union → ∩ | 0.99 | 0.59 | 0.74 | 236 | 2 | 167 |
| Intersection → ∪ | 0.97 | 0.85 | 0.90 | 341 | 12 | 62 |
| Intersection → ∩ | 0.99 | 0.51 | 0.68 | 206 | 1 | 197 |
| Strict-only → ∪ | 0.96 | 0.86 | 0.91 | 348 | 14 | 55 |
| Strict-only → ∩ | 0.99 | 0.53 | 0.69 | 212 | 1 | 191 |
| GPT fusion | SDoH subcategories | | | | | |
| | Precision | Recall | F1 | TP | FP | FN |
| Majority voting | 0.83 | 0.91 | 0.87 | 293 | 60 | 29 |
| Union | 0.70 | **<u>0.95</u>** | 0.80 | 305 | 131 | 17 |
| Intersection | **<u>0.91</u>** | 0.84 | **0.88** | 271 | 26 | 51 |
| Strict-only | 0.88 | 0.88 | **<u>0.88</u>** | 283 | 37 | 39 |

#### Supplementary Figures

Figure S1: Example annotation interface implemented in Label Studio.

Figure S2: Zero-shot prompt including task descriptions, SDoH definitions, and instructions.

Figure S3: 5-shot prompt including examples and accompanying explanations.

Figure S4: Forest plot of macro- and micro-averaged F1 scores with bootstrap 95% confidence intervals for domain category classification.

Figure S5: Forest plot of macro- and micro-averaged F1 scores with bootstrap 95% confidence intervals for subcategory classification.

**Figure S1.** Example annotation interface implemented in Label Studio.

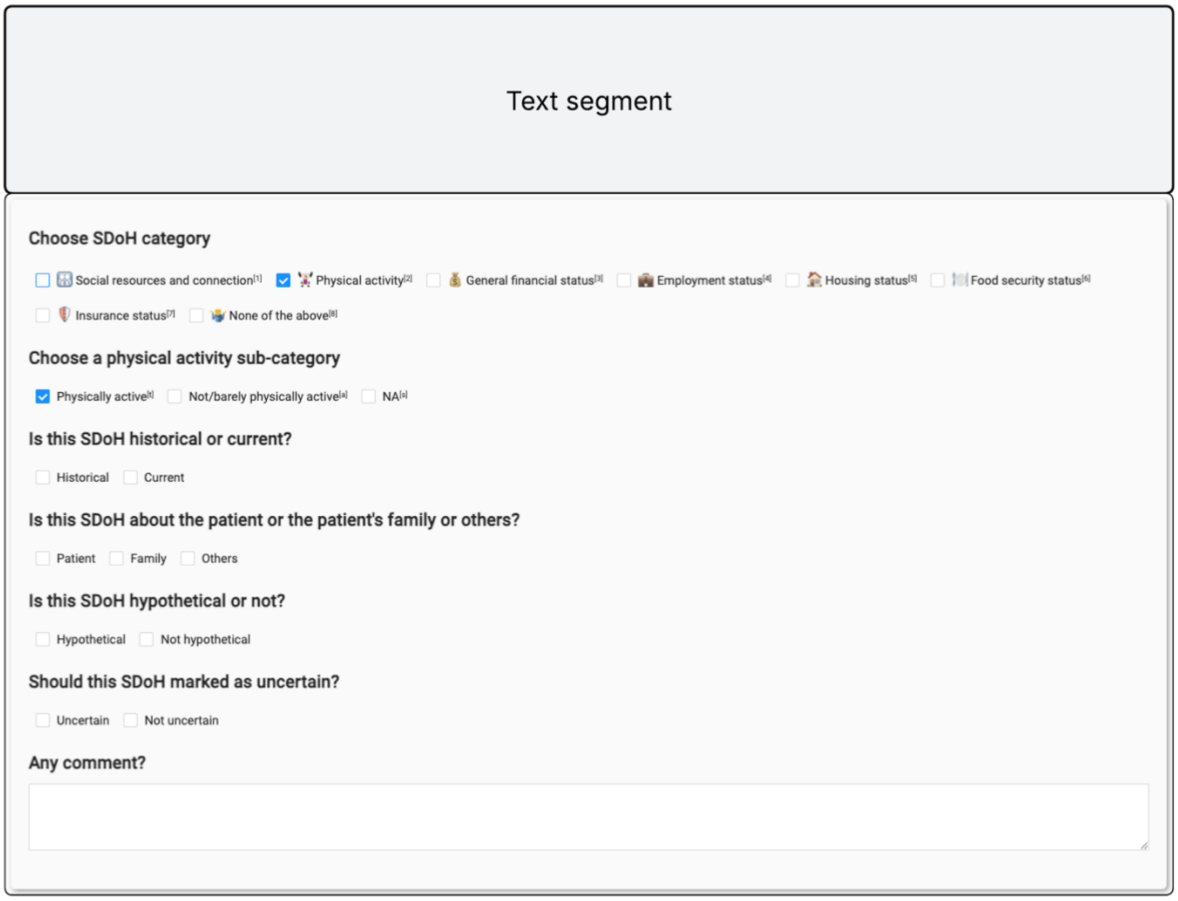

**Figure S2.** Zero-shot prompt divided into three sections: (1) task description; (2) definitions of SDoH domain categories and subcategories; and (3) instructions for SDoH extraction and output formatting.

SDoH Identification Prompt Structure (Zero-shot)

**Task role and description**

### Social Determinants of Health (SDoH) Identification Task
You're an expert social worker specialized in identifying social determinants of health (SDoH) in clinical notes. Your task is to analyze provided clinical text and identify relevant SDoH categories and sub-categories based ONLY on the explicit information in the text.

**SDoH domain categories, subcategories, and their definitions**

#### Predefined SDoH categories to identify (select all that apply or "none"):
{List of SDoH domain categories}

#### Predefined SDoH sub-categories to identify (select all that apply or "none"):
{List of SDoH subcategories}

#### SDoH definitions
*Physical activity*
Definition:
Physical activity (PA) refers to all movement including during leisure time, for transport to get to and from places, or as part of a person's work. Here PA includes both unstructured daily activities such as occupational or leisure time PA, and structured PA such as competitive sports.
Sub-categories:
*Physically active* - Participation in physical activities (light, moderate or vigorous) on a regular basis, ranging from walking to going to the gym.
*Not/barely physically active* - Physically inactive (or minimally active), or displays sedentary behavior.
... ...

**Instructions**

#### Instructions
1. Carefully read the clinical text chunk provided by the user.
2. Identify SDoH *categories* from the list above are explicitly present. Multiple selections are allowed.
  • If no relevant categories are mentioned, include only the string "none" in your categories list.
3. Identify SDoH *sub-categories* from the list above that correspond to the chosen categories. Multiple selections are allowed.
  • Each selected sub-category must belong to one of your selected categories.
  • If no relevant sub-categories are mentioned, include only the string "none" in your sub-categories list.
4. Only include the SDoH categories and sub-categories that meet ALL of the following criteria:
  • *Current* - Reflects the patient's present situation
    • Exclude events that are past conditions (e.g., "was homeless as a child") or resolved situations.
  • *Patient-specific* - Directly about the patient, not family members or others.
    • Exclude example: "Patient's brother lost his job".
  • *Actual* - Represents a real, observed status, not a plan, intention, or recommendation.
    • Exclude hypothetical scenarios, future plans, prospect, intentions, or recommendations, such as:
    • "Submitted a housing application" (future possibility)
    • "Pt is motivated and will attempt to be more active"
    • "If all goes well pt will likely move into a stable house"
    • "Recommended/discussed that pt increase physical activity"
  • *Confirmed* - Explicitly confirmed in the text, without uncertainty or pending status.
    • Exclude statements expressing uncertainty or ambiguity, such as:
      • SDoH question left unanswered (e.g., empty checkboxes) or marked "Pended"
      • "Pt is worried about possible eviction" (concern only)
      • "It may be the case that pt is homeless" (speculative)

**Output format**

5. Structure your response in JSON format using the `response_format` provided:
  • `SDoH_categories`: array of strings with identified categories or ["none"]
  • `SDoH_subcategories`: array of strings with identified sub-categories or ["none"]
6. Do **not** write any other text outside the JSON response.

(Think step-by-step internally if useful, but the user should see only the identified SDoH categories and sub-categories.)

**Figure S3.** 5-shot prompt structure, including examples and accompanying explanations.

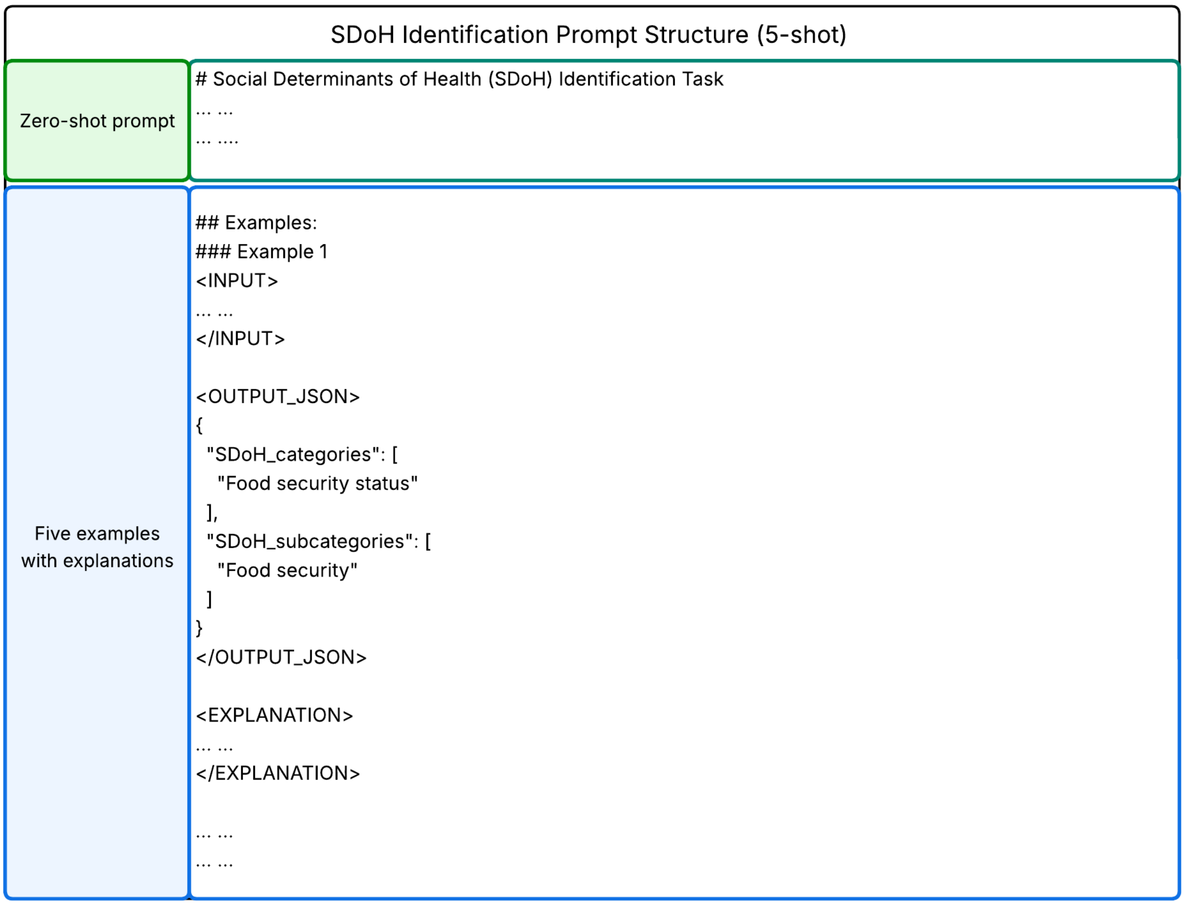

**Figure S4.** Forest plot of macro-averaged and micro-averaged F1 scores with bootstrap 95% confidence intervals for SDoH domain category classification.

Confidence intervals were computed using 2,000 patient-level cluster bootstrap resamples of the validation set.

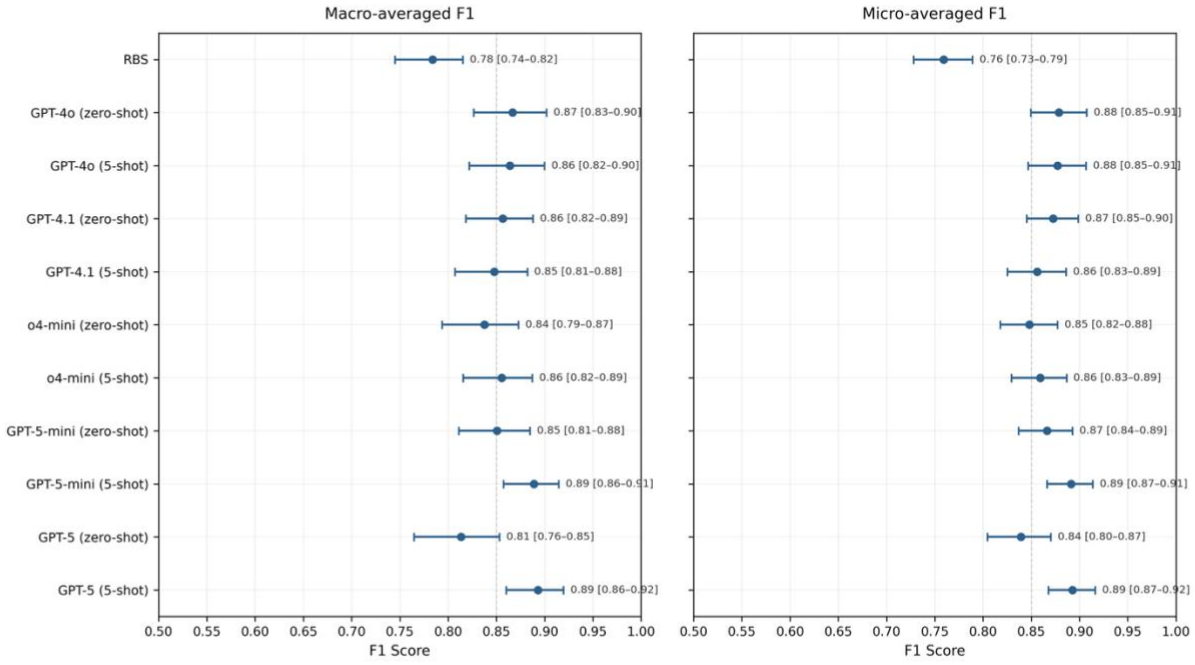

**Figure S5.** Forest plot of macro-averaged and micro-averaged F1 scores with bootstrap 95% confidence intervals for SDoH subcategory classification.

Confidence intervals were computed using 2,000 patient-level cluster bootstrap resamples of the validation set.

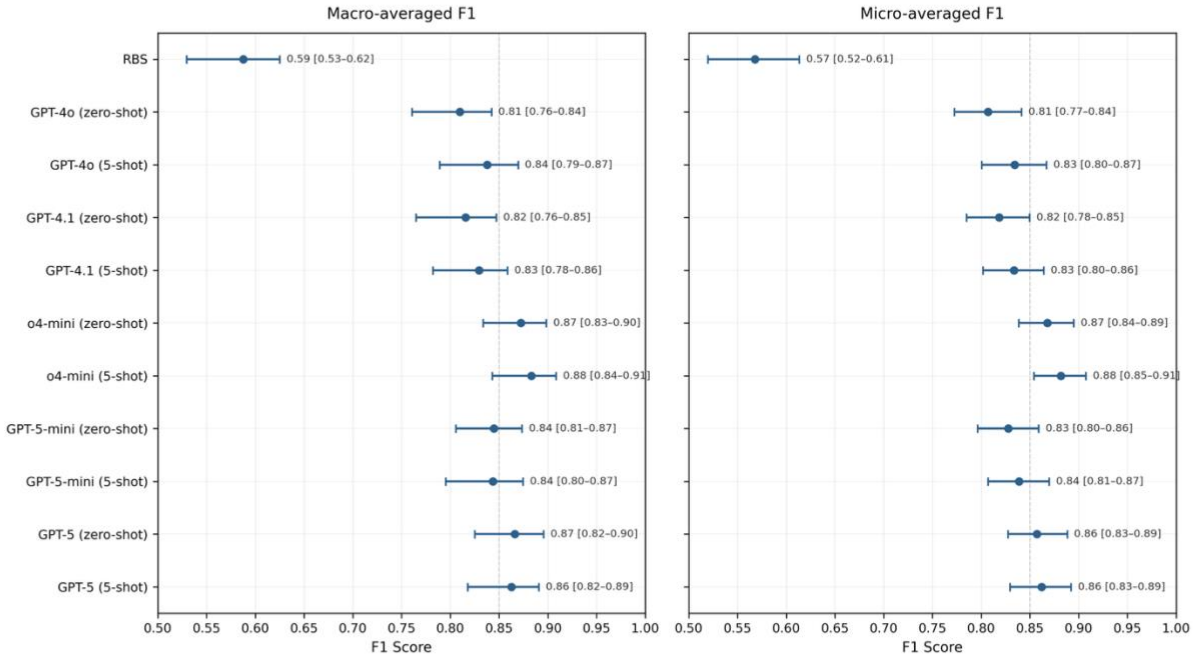
